# C4 genetic structural variations affect multiple sclerosis risk and progression

**DOI:** 10.1101/2025.07.28.25332292

**Authors:** Xin Lin, Dongdong Tang, Yuanhao Yang, Maria Pia Campagna, Melissa M. Gresle, Wei Z. Yeh, Sandeep Sampangi, Pavlina Kleinova, Fuencisla Matesanz, Antonio Alcina, Sara Eichau, Marzena J. Fabis-Pedrini, Mark Slee, Allan G. Kermode, Trevor Kilpatrick, Jeannette Lechner-Scott, Eva Kubala Havrdova, Dana Horakova, ANZgene Consortium, Helmut Butzkueven, Bruce V. Taylor, Vilija G. Jokubaitis, Yuan Zhou

## Abstract

**Objective:** Major histocompatibility complex (MHC) locus carries a significant genetic risk burden for multiple sclerosis (MS). Here we investigate the structurally diverse complement component 4 (*C4*) alleles within the MHC in MS development and disease progression.

**Methods:** We imputed and examined *C4* alleles based on data from two case-control cohorts (N_1_= 3252 cases and 5725 controls; N_2_= 8978 cases and 6976 controls), a clinical MS cohort (N_3_= 2387 cases) and a cohort with immune cell gene expression data (N_4_= 33 cases and 33 controls). We have performed gene-level analysis to investigate the shared genetic landscape between MS susceptibility (N= 14802 cases and 26703 controls) and plasma C4 protein (N= 68716).

**Results:** Our data showed that *C4* genetic structural variants were associated with significant changes in MS susceptibility and disability progression. For instance, higher *C4AL* copy number burden was associated with lower MS susceptibility (fixed effect meta-analysis odds ratio= 0.89, P= 5.65×10^−6^) independent of established MHC risk variants such as *HLA-DRB1*\*15:01. Higher C4AL copy number was also associated with reduced hazard of reaching MS disability milestones such as Expanded Disability Status Scale 3 (hazard ratio= 0.79, P= 9.0×10^−15^). In addition, we found *C4* alleles may also modulate *C4* expression in disease-relevant immune cell types such as CD8+ T cells. Further, we identified that candidate genes shared between MS susceptibility and plasma C4 protein level were enriched in biological pathways of immune regulation, Epstein-Barr virus infection and other autoimmune diseases such as lupus.

**Interpretation:** These findings support future investigations of the C4 genetic structural variants as potential mechanistic and therapeutic targets in MS pathogenesis and disease progression.

## INTRODUCTION

Multiple sclerosis (MS) is a disease of the central nervous system, characterised by a pathophysiological triad encompassing inflammation, demyelination, and axonal degeneration.^1^ MS affects approximately 2.8 million individuals worldwide, yielding a prevalence rate of 35.9 cases per 100,000 as of the year 2020.^2^ Large genome-wide association studies (GWAS)^3^ have established a significant genetic risk component for MS, including the most significant genetic risk loci (*HLA-DRB1*\*15:01) in the MHC^3^ as an important contributor to MS genetic susceptibility.

Complement activation has been implicated as a critical driver of neuroinflammation in neurodegenerative diseases such as Alzheimer’s.^4^ Located within the MHC Class III region, complement component 4 (C4) is involved in the classical and lectin pathways of complement activation that modulate a range of immune responses and T cell activation.^5,6^ The *C4* gene exists in two functionally distinct isoforms (*C4A* and *C4B*) that are distinguished by differences encoded by exon 26 (**Figure 1A**) and produce proteins binding to different molecular targets. Each of the isoforms further segregate in long (L) and short (S) forms (*C4AL, C4AS, C4BL, C4BS*) based on the presence of a human endogenous retrovirus (HERV) insertion at intron 9 without changing the C4 protein sequence.^7^ Emerging evidence supports that genetic structural variation (e.g., copy number ranging from one to three copies per haplotype) of the *C4* gene is highly relevant in neuropsychiatric diseases such as schizophrenia^8^ as well as autoimmune diseases including systemic lupus erythematosus (SLE) and Sjögren’s syndrome.^9^

**Figure 1.**
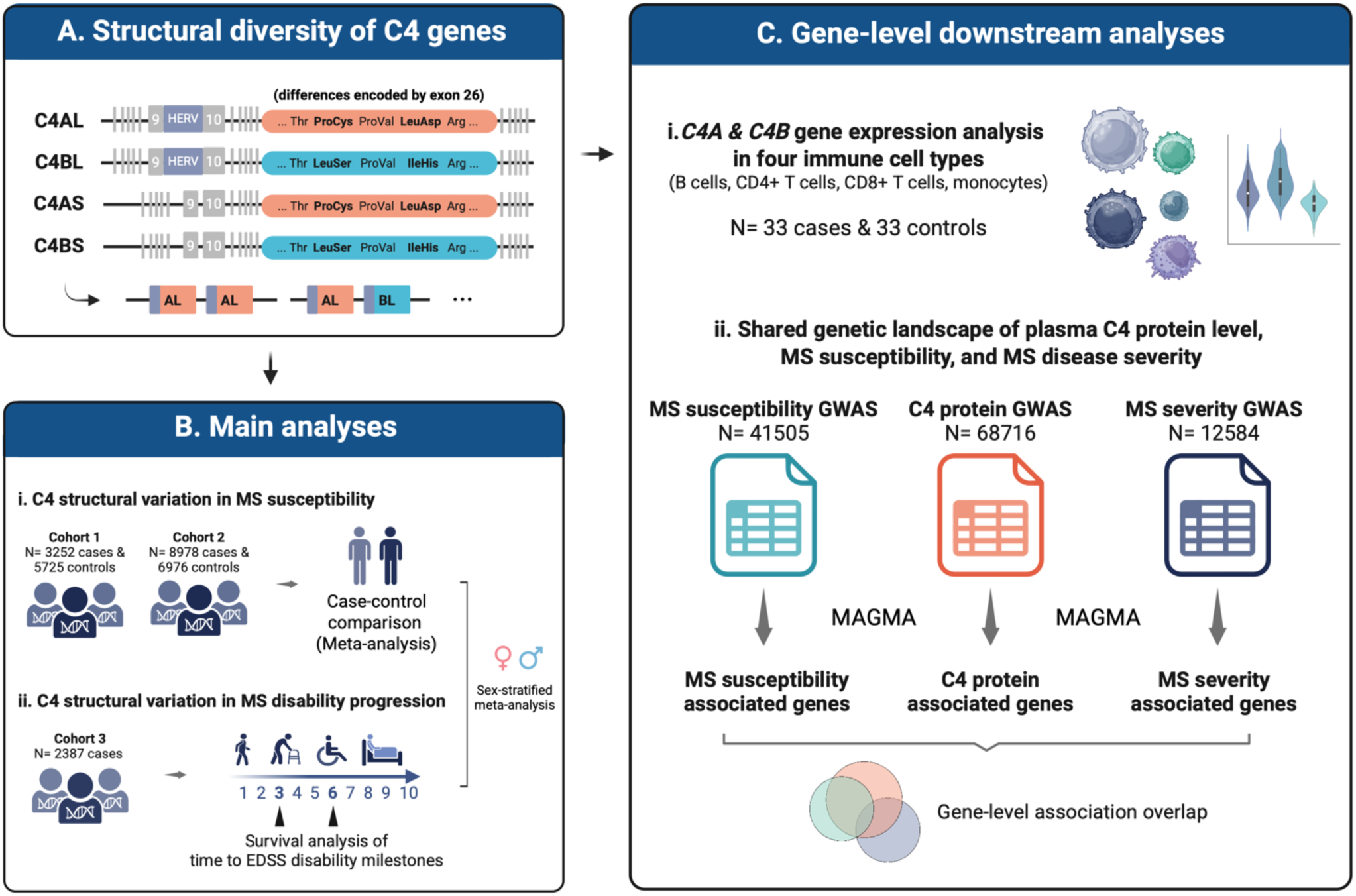
Study overview. **(A)** As described by Sekar et al.^7^, the C4A and C4B isoforms are distinguished based on differences encoded by exon 26. These isoforms could be further classified into long form (C4AL and C4BL) and short form (C4AS and C4BS) based on the presence of human endogenous retrovirus (“HERV”) between exon 9 and 10; **(B)** Main analyses conducted to assess associations of *C4* gene isoform copy number burden, *C4* structural haplotypes and diplotypes with: i. MS susceptibility; and ii. time to reaching MS clinical disability milestones (EDSS3 and EDSS6); **(C)** Gene-level downstream analyses included: i. *C4A* and *C4B* gene expression level across four immune cell types; and ii. investigating the shared genetic landscape of plasma C4 protein level, MS susceptibility, and MS disease severity. C4 = complement component 4. EDSS = expanded disability status scale. MAGMA = the multi-marker analysis of genomic annotation method. MS = multiple sclerosis.

A recent study found that copy number variation at the *C4* locus was associated with MS susceptibility.^10^ However, the study was based on limited sample size when compared to previous investigations in other diseases.^8,9^ Further, no study to date has evaluated the role of *C4* genetic structural variation in MS disease progression, and whether specific *C4* structural haplotype could be a potential genetic trigger and/or driver of disease activities in MS. Additionally, the functional consequences of *C4* genetic structural variation remain unclear, such as their potential impact on *C4* gene expression level. An improved understanding of the role of C4 in MS risk and progression may facilitate future studies of immunomodulatory therapies targeting the complement cascade. To address the knowledge gap, this study first systematically investigated the effects of *C4* genetic structural variants on the MS susceptibility and disability progression, including overall *C4* copy number burden, *C4* haplotypes and diplotypes. Further, we also examined the relationship between *C4* structural variation with *C4* gene expression level across different relevant immune cell types by leveraging data from an independent case-control cohort. To investigate the shared genetic landscape of C4 and MS, we then performed gene-level analysis to identify candidate genes and biological pathways associated with both C4 protein level and risk of MS, which present key targets for future studies of disease mechanisms and drug development.

## MATERIALS AND METHODS

As illustrated in **Figure 1**, this study employs several individual-level and summary-level datasets. Ethics approval and written informed consent were obtained in accordance with the Declaration of Helsinki.

### Individual-level data 1: MS cohort 1

The first study cohort encompassed genetic data from a total of 8977 unrelated individuals (n= 3252 MS cases and 5725 controls) with methodology described in detail previously.^11^ All cases and controls provided written informed consent in accordance with approval from appropriate local institutional ethics committees or institutional review boards as detailed in original investigations^12–15^. Genotyping was performed using Illumina Infinium Hap370CNV array for samples from the ANZgene Consortium, while Illumina Human660-Quad chip was used for samples as part of the GWAS study by the International MS Genetics Consortium (IMSGC) and the Wellcome Trust Case Control Consortium-2 (WTCCC2). The control samples from the Queensland Institute of Medical Research were genotyped on the Illumina Human610-Quad chip, and the Sentrix® HumanHap550 BeadChip was used for samples from the GeneMSA study. Quality control steps on the different datasets prior to merging as well as on the combined data, included SNP quality controls to remove SNPs with low call rates (less than 0.95) or in Hardy-Weinberg disequilibrium (p< 10^−7^), and per-sample filtering and principal component analysis using PLINK to exclude samples with low call rates, duplicates, close relatives and ancestry outliers.^11^ No further quality control was performed in this study.

### Individual-level data 2: MS cohort 2

The second study cohort was the WTCCC2 study cohort^13^ in which we obtained the individual-level genotyping data generated using the Illumina Human 660-Quad platform. As described previously,^13^ all individuals in this study provided informed consent in accordance with approval from the relevant local Ethical Committees or Institutional Review Boards. The original investigation^13^ applied quality control measures including per-sample genotyping quality control of call rates and heterozygosity rates, identity by descent analysis using a hidden Markov model to exclude related individuals, and principal component analysis clustering using *SHELLFISH* to exclude non-European ancestry samples. Additionally, stringent SNP quality controls were performed using a clustering algorithm and established pipelines detailed in the original publication. In this study, after removing samples overlapping with the MS cohort 1 dataset, we retained a total of 15954 unrelated individuals, including 8978 MS cases and 6976 controls.

### Individual-level data 3: immune cell C4A & C4B gene expression

The third cohort included a total of 66 unrelated individuals of European ancestry (33 relapsing-remitting MS cases and 33 healthy controls without any medical history of neurological or autoimmune disease) as previously described.^16^ Briefly, these participants were recruited from Royal Melbourne Hospital and Box Hill Hospital in Victoria, Australia from 2014 to 2016. With written informed consent obtained from all participants, this study was conducted according to the Declaration of Helsinki principles and was approved by the Human Research Ethics Committees of Royal Melbourne Hospital and Box Hill Hospital, Victoria, Australia.^16^ The MS cases were not on immunomodulatory treatment at the time of sample collection, and the clinical diagnosis was made by a neurologist and met the 2010 McDonald diagnostic criteria.^17^ Peripheral immune cells were isolated by cell sorting methods (Miltenyi Biotech and Stemcell Technologies). Only samples with greater than 90% purity of isolated immune cell subsets were included, which was assessed using standard flow cytometry protocols on a Cyan Flow cytometer (Beckman Coulter). RNA samples were prepared from the immune subsets using Qiagen RNeasy Mini Kit based on manufacturer’s protocol with DNase digestion. RNA sequencing was performed at the Australian Genome Research Facility using Illumina HiSeq 2500 system for the B cell and monocyte samples (approximately 40 million reads per samples generated) and Illumina Novaseq 6000 system for the CD4+ and CD8+ T cell samples (approximately 30-40 million reads per samples generated). Gene expression data were processed using edgeR package^18^ and RUVg method^19^ as established in previous work.^16^ No additional quality control was performed in this study.

### Individual-level data 4: longitudinal MS cohort

The fifth cohort with individual-level data is a longitudinal MS cohort^20^ genotyped using the Illumina MegaEx BeadChip array, as well as an additional customised panel of 3000 SNPs including: known multiple sclerosis risk SNPs, tag SNPs for classical HLA alleles, and previously published putative severity SNPs. The longitudinal cohort consisted of people with MS of European ancestry, which were genotyped in three tranches at the John. P. Hussman Institute for Human Genomics (University of Miami, USA). As described previously,^20^ all participants in this study gave written informed consent in accordance with approval by the Melbourne Health Human Research Ethics Committee and by institutional review boards at all participating centres. Clinical assessments were conducted every six months and entered by neurologists to collect clinical information such as disease-modifying therapy use, disease phenotypes, relapse information, and the Expanded Disability Status Scale (EDSS; an established measure of MS disability^21^). In this study, data from 2387 individuals were retained after extensive quality control steps per tranche as well as in the combined dataset, including principal component analysis to exclude outlier samples and per-SNP quality controls using PLINK 1.90. Our survival analysis of time to reaching EDSS3 disability milestone included only individuals with three or more visits and had not reached EDSS3 at the first recorded visit (n=1504). Additionally, a subgroup analysis was performed for individuals who had at least three visits, had not reached EDSS3 at first recorded visit, and subsequently reaching both EDSS3 and EDSS6 milestones during follow-up (n= 190).

### Individual-level data imputation

Prior to C4 imputation, all individual-level datasets underwent genotype imputation using the Michigan Imputation Server (Minimac4 version 1.7.3) based on the Haplotype Reference Consortium (HRC r1.1) reference panel (hg19) and phasing using Beagle version 5.4. Furthermore, for all individual-level datasets except the longitudinal MS cohort with custom genotyped HLA-tagging SNPs, we used the Michigan Imputation Server to perform imputation of the Human Leukocyte Antigen (HLA) region based on the Four-digit Multi-ethnic HLA version 1 (2021) reference panel and phasing using Eagle version 2.4. The post-imputation quality control measures included filtering for imputation quality (r^2^ > 0.2) and a MAF greater than 0.05. After these steps, C4 allele imputation from MHC genotypes was then performed using the established *imputec4* protocol.^8^ Similar to the previous study,^8^ frequencies of imputed C4 alleles are highly consistent between our study cohorts as illustrated in **Figure 2A** (C4 copy numbers), **Figure 3A** (C4 haplotypes) and **Supplementary Figure S1A** (C4 diplotypes).

**Figure 2.**
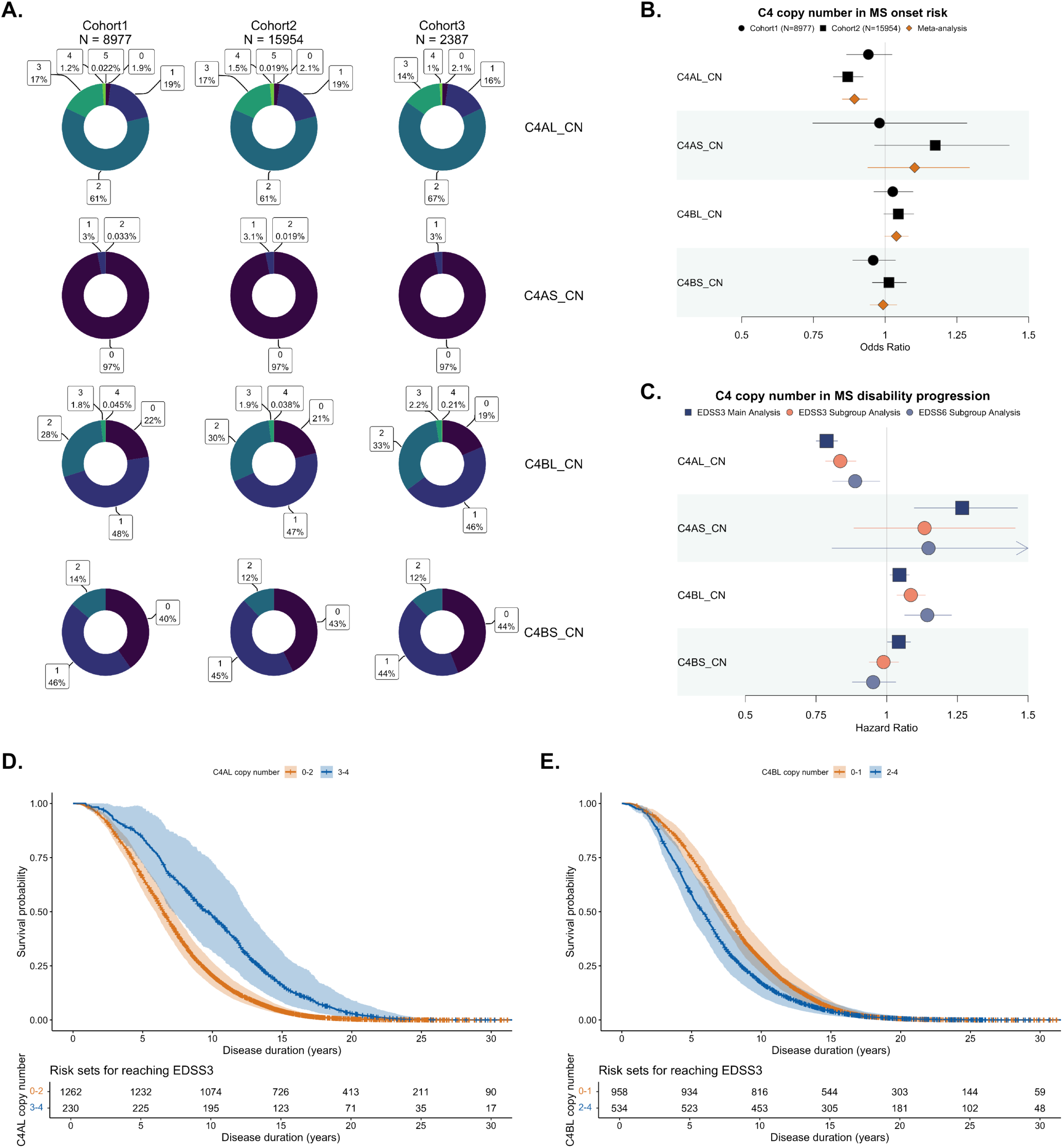
*C4* copy number associations with MS susceptibility and disability progression. **(A)** Frequency map of *C4* copy numbers across three study cohorts; **(B)** Meta-analysis of *C4* copy number associations with MS susceptibility, which were adjusted for sex, top 10 PCs and *HLA-DRB1*\*15:01; **(C)** *C4* copy number associations with risk of MS disability progression, adjusted for age at onset, sex, top 10 PCs and *HLA-DRB1*\*15:01 tagging SNP (rs3135388); **(D-E)** Kaplan-Meier plots of survival probability for reaching EDSS3 disability milestones in main analysis of study cohort 3 by different copy numbers of C4AL and C4BL. C4 = complement component 4. C4AL_CN = copy number of C4AL. C4BL_CN = copy number of C4BL. C4AS_CN = copy number of C4AS. C4BS_CN = copy number of C4BS. EDSS = expanded disability status scale.

**Figure 3.**
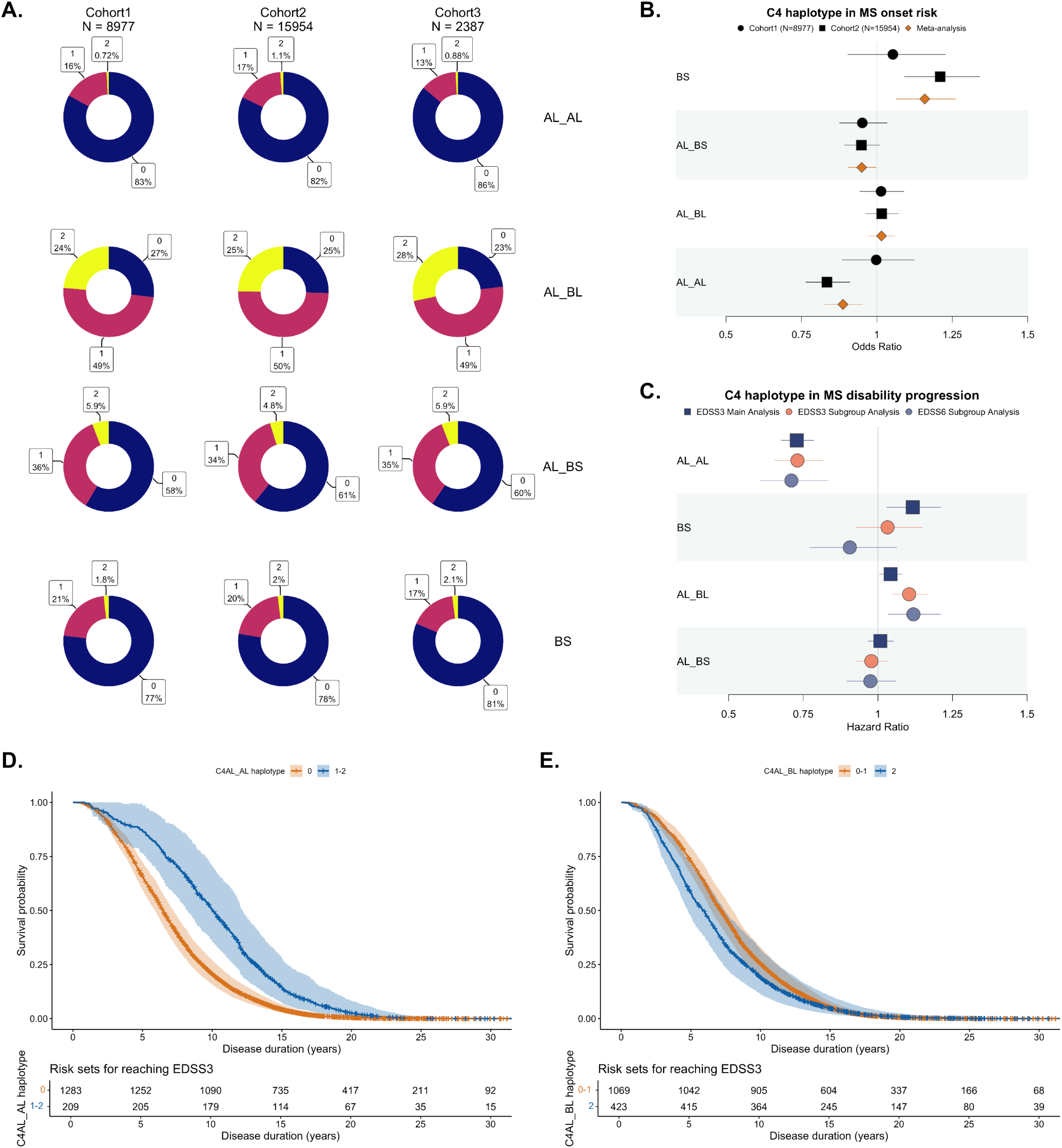
*C4* haplotype associations with MS susceptibility and disability progression. **(A)** Frequency map of *C4* haplotypes across three study cohorts; **(B)** Meta-analysis of *C4* haplotype associations with MS susceptibility, which were adjusted for sex, top 10 PCs and *HLA-DRB1*\*15:01; **(C)** *C4* haplotype associations with risk of MS disability progression, adjusted for age at onset, sex, top 10 PCs and *HLA-DRB1*\*15:01 tagging SNP (rs3135388); **(D-E)** Kaplan-Meier plots of survival probability for reaching EDSS3 disability milestones in main analysis of study cohort 3 by AL_AL and AL_BL haplotypes. C4 = complement component 4. EDSS = expanded disability status scale.

### Summary-level data 1: MS susceptibility GWAS

The GWAS summary dataset pertaining to MS susceptibility was acquired from a comprehensive meta-analysis conducted by the International MS Genetics Consortium (IMSGC).^3^ This analysis amalgamated data from 15 distinct datasets, encompassing 14802 MS cases and 26703 controls, all European descent. We annotated allele frequencies for SNPs using the 1000 Genome Project phase 3 EUR population reference,^22^ and excluded SNPs that were either multiallelic, ambiguous, with minor allele frequency <0.05, or not present in all 15 GWAS cohorts. After these steps, a total of 5.5 million SNPs were retained for subsequent analysis.

### Summary-level data 2: MS severity GWAS

The MS severity summary data^23^ was generated from 21 cohorts collected from various centers in North America, Europe, and Australia. This dataset focused on a genetic analysis of disease severity in 12584 individuals with MS of European ancestry. MS disease severity was quantified using the cross-sectional age-related MS severity (ARMSS) score, an established numerical scale that correlates with the progression of neurological degeneration. Following quality control measures, approximately 6.6 million SNPs were retained for subsequent analysis.

### Summary-level data 3: C4 protein GWAS

The C4 protein GWAS summary dataset was derived from the iPSYCH 2012 case-cohort study of 68768 neonatal samples.^24^ Briefly, specific antibodies (MA1-72520 (ThermoFisher Scientific) and HYB162-04) was used to measure total C4 protein concentrations, and the samples were genotyped using the Infinium PsychChip v1.0 array (Illumina, San Diego, CA, USA) at the Broad Institute (Boston, MA, USA). Post-quality control, the dataset was consolidated to approximately 4.5 million SNPs for analysis.

### C4 association analysis

We used the “glm” function within R software (version 4.0.3) to evaluate the associations of *C4* copy number burden and the structurally diverse *C4* alleles with MS susceptibility. In the multivariable models, we adjusted for the top 10 genome-wide principal components (PCs), sex, and *HLA-DRB1*\*15:01 as covariates. Furthermore, a fixed-effect meta-analysis was conducted on the two case-control cohorts (Cohort 1 and Cohort 2) using the “meta” R package (version 8.0-1). Further, we conducted survival analysis based on Cox proportional hazard model to assess associations of *C4* genetic structural variants with time to reaching EDSS disability milestones. Our haplotype analyses focused on the four most prevalent haplotypes (AL_BL, AL_BS, BS, and AL_AL; see **Figure 3A).** All analyses of C4 diplotypes focused on diplotypes with at least 5% frequency and we used the diplotype with highest frequency as the reference group. Following the analyses of C4 associations with MS susceptibility and disability progression, we performed sex-stratified analyses to assess potential sex-biased or sex-specific associations. As a sensitivity analysis, we utilized the “glmnet” R package (version 4.1-8) to perform Lasso regression after removing covariates that had moderate correlation with *C4* copy number or *C4* haplotype (Pearson correlation r> 0.5). For the analysis of *C4A* and *C4B* expression level across different immune cell types, the top 10 genome-wide PCs and sex were used as covariates in multivariable model.

### MAGMA and KEGG enrichment analysis

We implemented MAGMA^25^ to examine gene-level associations based on established protocols and used a window size of 10kb upstream & 1.5kb downstream for mapping SNPs to genes. For the plasma C4 protein GWAS MAGMA analysis, we applied the False Discovery Rate (FDR) correction method^26^ to account for multiple comparisons, setting a significance threshold at P_FDR_< 0.05. Based on the identified candidate genes associated with the plasma C4 protein levels, we then employed a significance threshold of P< 0.05 to assess overlapping genes between MS susceptibility, MS severity, and plasma C4 protein levels. Using the candidate genes identified from MAGMA, we then performed KEGG pathway enrichment analysis using the clusterProfiler R package (v4.12.6)^27^ with an FDR-adjusted p-value threshold of 0.05 and a q-value cutoff of 0.02.

## RESULTS

### Copy numbers of C4 genes are associated with MS susceptibility and disability progression

A meta-analysis of the two case-control cohorts identified an association of higher *C4AL* copy number with lower MS susceptibility, persisting on adjustment for sex, the top 10 PCs, and the *HLA-DRB1*\*15:01 allele (OR= 0.89, P= 5.65×10^−6^; see **Figure 2B** and **Supplementary Table 1**). After removing covariates that had moderate correlation with *C4AL* copy number (PC2 in Cohort1; **Supplementary Figure S2**), the sensitivity analysis revealed concordant estimated direction of effects for *C4AL* copy number across both study cohorts (**Supplementary Table 2**). To further assess the robustness and independence of these associations, we conducted additional sensitivity analyses adjusting for independent MHC SNPs identified in previous MS susceptibility GWAS.^3^ We found our key findings including the *C4AL* copy number associations attenuated but persisted upon adjustment (P< 2.19×10^−4^; **Supplementary Table 3**), further supporting that these are independent association signals in the MHC region for MS.

In an independent clinical cohort, survival analysis found that higher copy numbers of *C4AL* were associated with reduced hazard of reaching the EDSS3 disability milestone, which persisted on adjustments (HR= 0.79, P= 9.0×10^−15^; **Figure 2C-D**; **Supplementary Table 4**). In the subgroup analysis of individuals who reached both EDSS3 and EDSS6 during follow-up (illustrated in **Figure 2C**), we observed consistent association patterns where higher copy numbers of *C4AL* were associated with reduced hazards of reaching EDSS3 (HR= 0.84, P= 5.1×10^−8^) and EDSS6 (HR= 0.89, P= 0.013). Further, we found higher copy numbers of *C4BL* were associated with increased hazards of reaching EDSS milestones in both the main analysis (EDSS3: HR= 1.05, P= 7.4×10^−3^) and subgroup analysis (EDSS3: HR= 1.09, P= 5.6×10^−4^; EDSS6: HR= 1.14, P= 2.8×10^−4^; **Figure 2C&E**; **Supplementary Table 4**). Additional sensitivity analyses also supported that the key findings of *C4AL* and *C4BL* copy number associations with EDSS3 outcome persisted upon adjustments for clinical phenotype and DMT use as additional covariates (**Supplementary Table 4**). All main analysis findings of C4 copy numbers were robust on multiple testing correction by copy number types (P_threshold_ = 0.05/4 = 0.0125).

We also performed sex-stratified analyses (see **Supplementary Figure S3** and **Supplementary Tables 5 & 6**) and found similar association patterns of *C4AL* copy numbers in females and males where higher *C4AL* copy numbers were associated with reduced risk of both MS susceptibility (OR_male_= 0.91, P_male_= 0.015; OR_female_= 0.88, P_female_= 1.14×10^−4^) and disability progression (HR_male_= 0.71, P_male_= 1.70×10^−13^; HR_female_= 0.80, P_female_= 8.10×10^−14^). In contrast, there may be sex-biased effects of having higher *C4BL* copy number on increasing MS susceptibility in males (OR_male_= 1.07, P_male_= 0.03; OR_female_= 1.02, P_female_= 0.53) and risk of disability progression in females (HR_male_= 0.96, P_male_= 0.13; HR_female_= 1.09, P_female_= 3.10×10^−5^). We did not observe significant sex-specific effects of *C4* copy numbers acting in opposing directions for females and males (see **Supplementary Tables 5 & 6**).

### Structurally diverse C4 alleles are also implicated in MS susceptibility and disability progression

Next, we assessed whether specific structural haplotypes of the *C4* gene underlie the overall copy number associations. As shown in **Figure 3B** and **Supplementary Table 1**, our meta-analysis suggested the AL_AL haplotype (OR= 0.89, P= 9.49×10^−4^) was associated with reduced MS susceptibility, while the BS haplotype was associated with increased MS risk (OR= 1.16, P= 7.58×10^−4^). When considering haplotypes as categorical variables, similar results were observed (see **Supplementary Table 7**).

However, our sensitivity analysis did not find consistent effect estimates for BS haplotypes on MS susceptibility across two study cohorts (**Supplementary Table 2**).

Further, we found that the AL_AL haplotype was associated with longer time to reaching the EDSS3 disability milestone (HR= 0.73, P= 1.1×10^−16^; **Figure 3C-D**), while the AL_BL haplotype was associated with increased hazards of reaching EDSS3 (HR= 1.04, P= 0.027; **Figure 3C&E**) despite not persisting on multiple testing correction by tested C4 haplotypes (P_threshold_ = 0.05/4 = 0.0125). The subgroup analysis also revealed consistent association patterns where the AL_AL haplotype was associated with decreased hazard of EDSS3 (HR= 0.73, P= 3.3×10^−8^) and EDSS6 (HR= 0.71, P= 2.8×10^−5^; **Supplementary Table 3**). Further sensitivity analyses also supported that the key findings of AL_AL haplotype associations persisted upon adjustments for clinical phenotype and DMT use as additional covariates (**Supplementary Table 3**).

We additionally performed analyses of C4 diplotypes (i.e., combinations of C4 haplotypes; see **Supplementary Figure S1A-C and Supplementary Tables 8-9**) and found the AL_BL/AL_AL diplotype was inversely associated with hazard of reaching EDSS3 in reference to the most prevalent AL_BL/AL_BL diplotype (HR= 0.66, P= 1.3×10^−12^; **Supplementary Figure S1C-D and Supplementary Table 9**). Moreover, the AL_BL/AL_AL diplotype was also associated with reduced hazard of reaching EDSS3 (HR= 0.49, P= 1.2×10^−11^) and EDSS6 (HR =0.52, P =5.4×10^−5^; **Supplementary Table 9**) in subgroup analysis.

In the sex-stratified analysis, we found consistent direction of effects for AL_AL haplotype in females and males on MS susceptibility (OR_male_= 0.90, P_male_= 0.067; OR_female_= 0.88, P_female_= 0.006) and disability progression (HR_male_= 0.65, P_male_= 5.10×10^− 9^; HR_female_= 0.73, P_female_= 4.60×10^−12^). We did not observe significant sex-specific effects of *C4* haplotypes acting in opposing directions for females and males (see **Supplementary Figure S3** and **Supplementary Tables 4 & 5** for full results).

### C4 associations with differential C4A and C4B expression in immune cells

Our analysis of *C4* gene copy numbers and structural haplotypes identified associations with cell-type-specific *C4A* and *C4B* expression profiles (**Supplementary Figures S4-10**). We observed MS-specific effect of higher *C4AL* copy number associated with higher *C4A* expression in B cells (**Supplementary Figure S11**), but not for other *C4* copy number types and haplotypes when stratified by MS case-control status (**Supplementary Figures S12-16**). In CD8+ T cells, there was a positive association between increased *C4AL* copy number and increased overall *C4A* expression level robust on adjustments (β= 0.18, P= 0.024), and that AL_AL (β= 0.31, P= 0.039) and AL_BS (β= 0.25, P= 0.047) haplotypes both were also associated with higher C4A expression (see **Supplementary Table 10**). However, these did not pass multiple testing correction by immune cell types (P_threshold_ = 0.05/4 = 0.0125) likely due to limited study power from sample size. We also observed associations of *C4BL* copy number (β= -0.32, P= 1.90×10^−4^), AL_BS haplotype (β= 0.34, P= 0.009) and AL_BL haplotype (β= -0.28, P= 0.004) with *C4B* expression in CD8+ T cells (**Supplementary Table 11**). In monocytes, we found that higher *C4AL* copy numbers were associated with increased *C4A* expression (β= 0.14, P= 0.011; **Supplementary Table 10**).

### Shared genetic landscape between C4 and MS risk

Based on FDR-significant gene-level associations of C4 plasma protein level, we found 259 gene-level associations shared between C4 plasma protein and MS susceptibility and seven genes were associated with both C4 plasma protein level and MS severity (**Figure 4A; Supplementary Tables 12-13**). Among these identified candidates, five genes located within the MHC region emerged as shared across all three traits (*HLA-DRA*, *C4B*, *MAS1L*, *OR11A1*, and *OR12D2*; see **Supplementary Table 14**). Although *HLA-DRA* variants were associated with all three traits, this signal may also be attributable to linkage disequilibrium with the established MS risk allele *HLA-DRB1*\*15:01. Nevertheless, this supports the potential role of antigen presentation pathway in MS pathogenesis, as well as an interplay between adaptive immunity and complement-mediated neurodegeneration. Based on the candidate genes shared between C4 protein and MS susceptibility, we conducted KEGG pathway analysis and subsequently identified over-represented pathways as shown in **Figure 4B**. Many of the pathways were immune-related, including the top pathways such as antigen processing and presentation, Epstein-Barr virus (EBV) infection, and diseases such as SLE and inflammatory bowel disease (**Supplementary Table 15**). Example pathways with highlighted candidate genes have been illustrated in **Supplementary Figures S17-20**.

**Figure 4.**
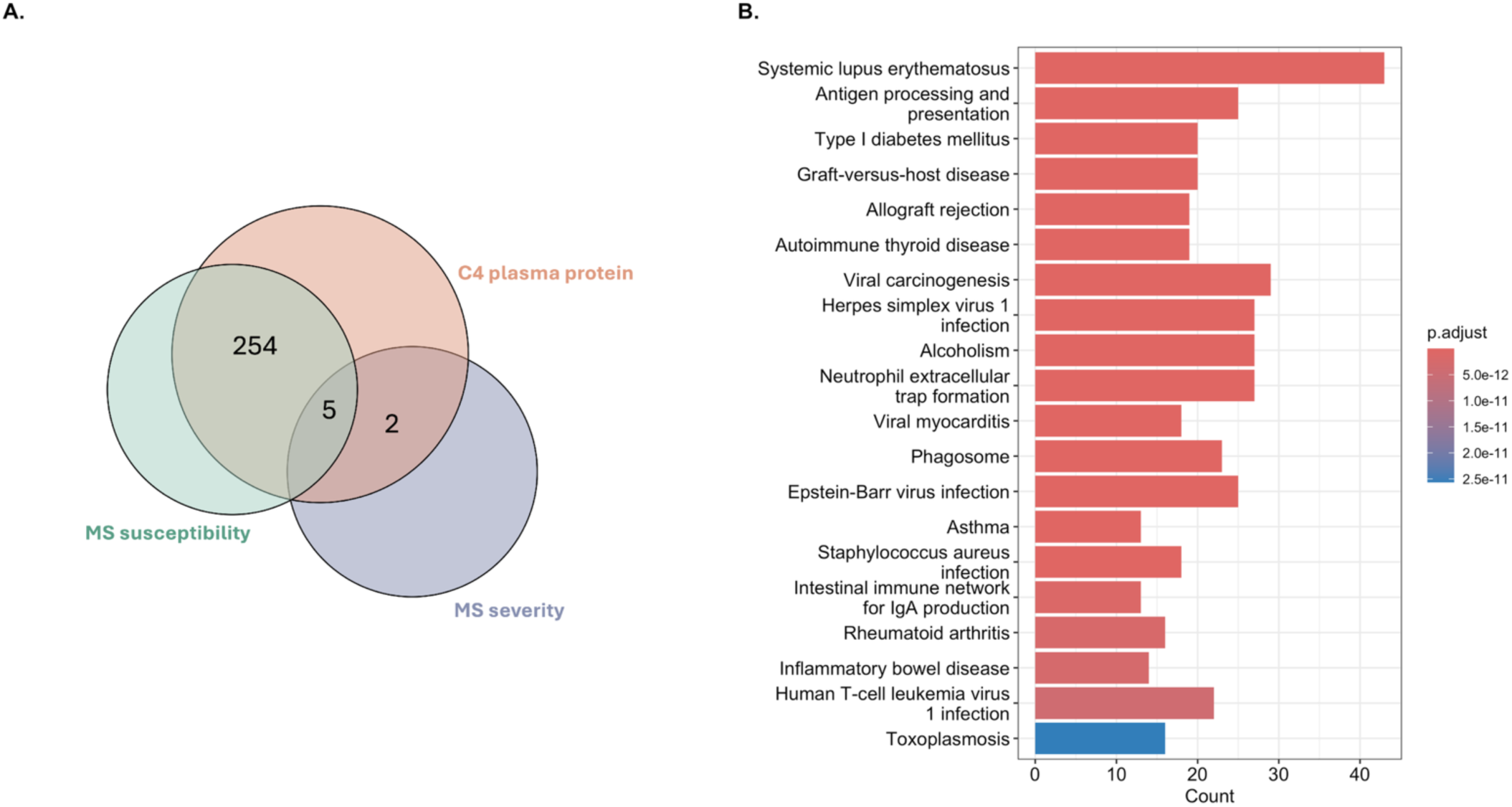
Shared genetic landscape of plasma C4 protein level, MS susceptibility and MS severity. **(A)** Based on FDR-significant gene-level associations for plasma C4 protein level (P_FDR_< 0.05) using the multi-marker analysis of genomic annotation method (MAGMA), there were 259 genes that were also associated with MS susceptibility (P< 0.05), seven genes with MS severity (P< 0.05) and five of the identified genes were shared across the three traits; **(B)** KEGG pathway over-representation analysis based on the 259 gene-level associations with plasma C4 protein level and MS susceptibility. C4 = complement component 4. Count = number of genes enriched in the pathway. p.adjust = adjusted p-value using the Benjamini-Hochberg procedure.

## DISCUSSION

C4 proteins encoded by the *C4* genes within the MHC locus on chromosome 6 are a pivotal part of the complement cascade, mediating both inflammation and neuronal damage.^28,29^ Our data shows that *C4* genetic structural variants are associated with significant changes in the risk of development of MS and the progression of MS. This is the largest study of *C4* genetic structural variation in MS to date, which adds critical insights into the MHC region as an important genetic risk locus for MS. We found that *C4* copy number burden, as well as specific *C4* haplotypes and *C4* diplotypes modulated both MS susceptibility and disability progression. In an independent case-control cohort, we further assessed the potential functional impact of *C4* alleles on gene expression across disease-relevant immune cell types such as CD8+ T cells, B cells and monocytes. Additionally, integration of large summary-level data enabled the identification of associated genes shared between C4 protein levels and MS risk, in which the identified genes were enriched in immune-related pathways including antigen processing and presentation, EBV infection, and autoimmune diseases. Together, these findings highlight a biologically plausible and important role of C4 in MS susceptibility and disease progression, which could lead to future studies of the complement cascade in MS and possibly developing new immunomodulatory drugs for targeting C4-related pathways.

As a crucial component of the complement system, the potential role of *C4* in MS has garnered considerable attention. The concentration of C4a (an activated fragment of C4) in plasma and cerebrospinal fluid of MS patients were significantly elevated compared to controls, with even higher levels observed during acute relapses.^30^ Further, previous studies of animal models have suggested the complement system as a double-edged sword in MS pathophysiology. For instance, complement proteins facilitate the clearance of myelin degradation products and other debris through myeloid cell-mediated phagocytosis, potentially aiding in tissue repair.^31^ However, complement deposition has been found in subtypes of MS white matter plaques supporting aberrant complement activation as a possible mechanism of exacerbating demyelination.^32^ Additionally, the complement system may interact with established risk factors for MS such as EBV infection, through the binding of complement fragments (e.g., C3d) and EBV gp350/220 to complement receptor 2 on the surface of B cells that may potentially lower the threshold of B cell activation and further sustain the activation of CNS self-reactive B cell populations.^31,33,34^ More recent findings also suggested an association of higher neonatal plasma C4 protein concentration with reduced risk of MS development^24^ and that overall higher *C4* copies conferred lower MS risk,^10^ which further support an important role of C4 in MS pathogenesis.

It has been long documented that C4A and C4B exhibit functionally distinct properties such as differences in binding reactivity to antigenic structures.^35,36^ It is thus biologically plausible that different genetic structural variants of the *C4* gene may have varying impacts on related disease susceptibility as evidenced in previous studies.^8,9^ Adding to existing literature, our findings based on multiple large study cohorts provide multiple lines of evidence for an important role of *C4* genetic structural variation (such as *C4AL* and *C4BL* copy numbers) in MS susceptibility and disability progression. Differential gene expression analysis in an independent study cohort additionally revealed associations of *C4AL* copy numbers as well as the AL_BS haplotype and the AL_AL haplotype with *C4A* expression in CD8+ T cells, supporting the hypothesis that *C4* alleles may have functional consequences that modulate expression levels of C4related genes. Furthermore, we examined the shared genetic architecture between plasma C4 protein level and MS risk, which led to the identification of candidate genes that are enriched in immune pathways. For example, the EBV infection pathway could be highly relevant since EBV infection is a well-established requisite environmental risk factor for MS development.^37,38^ Our results therefore warrant future studies of the complement cascade as a potential mechanism underpinning the role of EBV in modulating risk of MS. Moreover, previous genetic studies have reported associations of *C4* structural variation with SLE and Sjögren’s syndrome in similar direction of effects as our findings for MS.^9^ In addition, previous work has also identified a shared genetic architecture between inflammatory bowel diseases and MS.^39^ Given the emerging links between these diseases with MS, C4 genetic structural variation and the downstream functional consequences in the complement cascade might serve as a shared immunological pathogenic mechanism that warrants further investigations. Interestingly, current complement drug development are targeting complement cascade molecules other than C4, including Eculizumab which targets C5 and has been shown to be effective in neuromyelitis optica^40^ (clinical trial ID: NCT04355494) and the preclinical phase PMX205 targeting the C5a receptor suggestive for neurodegenerative disease such as Alzheimer’s disease.^41^ As our understanding of the complement system in human diseases continues to evolve, future research and therapeutic development may benefit from targeting C4.

This study has several strengths that underscore the robustness and reliability of our findings. First, the ample statistical power derived from large sample sizes significantly facilitates the detection of associations, enhancing the validity of our results. Next, the integration of a diverse range of genomic, transcriptomic, and clinical data provides a comprehensive, multi-layered perspective on the role of C4 in MS, enriching our understanding through multidimensional insights. However, it is essential to acknowledge the limitations inherent in this study. For instance, the MS case cohorts may include individuals with comorbidities (e.g., SLE) that had also been associated with C4 genetic structural variants, where we could not perform a sensitivity analysis due to lack of data availability. Further, another limitation of this study would be that the C4 structural variation was imputed from genotyping array data rather than direct genotyping or sequencing, though the imputation accuracy was reasonably high (0.70 < r^2^ < 1.00) for the four most common structural forms of the *C4A/C4B* locus (BS, AL_BS, AL_BL, and AL_AL).^8^ An important future direction would be generating better imputation reference data validated for different genotyping array types by direct sequencing large population cohorts with diverse ancestry backgrounds. In addition, findings from our exploratory analyses in the longitudinal cohort supported further validation studies of C4 in MS disease progression, as well as further mechanistic studies utilising animal and cellular models of neuroinflammation. We also observed B-cell C4A expression effect only in MS cases in our exploratory cell-specific analysis, though this may reflect limited sample size and require further validation from other study cohorts in future studies. This is an important future direction as B cells are the main targets of EBV infection and could contribute to MS pathophysiology via potential mechanisms such as antibody production and antigen presentation. Furthermore, C4 structural variation, gene expression, and protein levels have not been measured and assessed within the same individuals. Future studies could benefit from direct sequencing coupled with transcriptomic and proteomic profiling in the same study cohorts to perform a more comprehensive and integrated analysis of C4 genetic structural variants. Additionally, the relatively smaller sample size for gene expression data could constrain the statistical power for detecting subtle expression differences.

## CONCLUSION

Out study highlights a significant role of *C4* genetic structural variation in MS susceptibility and disability progression, possibly through modulating *C4* gene expression in immune cells (e.g., CD8+ T cells) as well as genetic regulation of immune responses. In the shared genetic landscape between C4 protein levels and MS susceptibility, we identified key candidate genes that were enriched in biological pathways regulating immune responses, EBV infection, and several other immune-related diseases. Together, these findings hold significant implications for an improved understanding of the genetic triggers and drivers of MS, while providing valuable insights to support future mechanistic studies and potentially therapeutic development for MS.

## ACKNOWLEDGEMENT

We thank the research participants whose involvement make this work possible, as well as the investigators who have made their datasets available to undertake research of this type. We would also like to acknowledge the use of the high-performance computing facilities provided by Digital Research at the University of Tasmania, as well as by eResearch high-performance computing team at the Monash University. This work was supported by funding from Multiple Sclerosis Australia (XL: 23-PDF-0120), the Excellent Young Backbone Talents Overseas Visiting and Training Program of Anhui Medical University (DT: gxgwfx2020025), the Mater Foundation (YY), Agencia Española de Investigación (AEI)-FEDER (FM, AA: RED2022-134425-T and PID2022-138400OB-C21), and the Australian National Health and Medical Research Council (BVT: 2009389; YZ: 1173155).

## AUTHOR CONTRIBUTIONS

DT, XL, and YZ contributed to conceptualisation and visualisation; DT, XL, YY, and YZ contributed to methodology; MPC, MMG, WZY, SS, PK, FM, AA, SE, MJFP, MS, AGK, TK, JLS, EKH, DH, HB, BVT, VGJ, YZ, and the ANZgene Consortium contributed to data curation; DT and XL contributed to the formal analysis; BVT, VGJ, and YZ contributed to supervision; DT, XL, BVT, and YZ contributed to funding acquisition; DT, XL, YY, BVT, VGJ, and YZ contributed to original draft manuscript; DT, XL, YY, MPC, MMG, WZY, SS, PK, FM, AA, SE, MJFP, MS, AGK, JLS, EKH, DH, HB, BVT, VGJ, and YZ contributed to review and editing of manuscript.

### The members of the ANZgene Consortium are

Alan Baxter (School of Pharmacy and Molecular Sciences, James Cook University, Townsville, Australia), Allan G. Kermode (Department of Neurology, Sir Charles Gairdner Hospital, Nedlands, Australia), Melanie Bahlo (Bioinformatics Division, Walter and Eliza Hall Institute of Medical Research, Parkville, Australia), William Carroll (Department of Neurology, Sir Charles Gairdner Hospital, Nedlands, Australia), Helmut Butzkueven (Department of Medicine, Royal Melbourne Hospital, Parkville, Australia), David Booth (Westmead Millennium Institute, University of Sydney, Sydney, Australia), Graeme Stewart (Westmead Millennium Institute, University of Sydney, Sydney, Australia), James Wiley (Howard Florey Institute, University of Melbourne, Melbourne, Australia), Judith Field (Howard Florey Institute, University of Melbourne, Melbourne, Australia), Lotti Tajouri (Genomics Research Centre, Griffith University, Brisbane, Australia), Lyn Griffiths (Genomics Research Centre, Griffith University, Brisbane, Australia), Michael Barnett (Brain and Mind Research Institute, University of Sydney, Camperdown, Australia), Pablo Moscato (Hunter Medical Research Institute, Newcastle, Australia), Robert Heard (Westmead Millennium Institute, University of Sydney, Sydney, Australia), Rodney Scott (School of Biomedical Sciences, University of Newcastle, Newcastle, Australia), Shaun McColl (School of Molecular & Biomedical Science, University of Adelaide, Adelaide, Australia), Simon Foote (Australian School of Advanced Medicine, Macquarie University, Sydney, Australia), Simon A Broadley (School of Medicine and Dentistry, Griffith University, Gold Coast Campus, Australia), Mark Slee (School of Medicine, Flinders University of South Australia, Adelaide, Australia), Steve Vucic (Western Clinical School, University of Sydney, Randwick, Australia), Trevor Kilpatrick (Centre for Neurosciences, Department of Anatomy and Neuroscience, University of Melbourne, Melbourne, Australia), Justin Rubio (The Florey Institute of Neuroscience and Mental Health, Melbourne, Victoria, Australia), Allan Motyer (School of Mathematics and Statistics, The University of Melbourne, Melbourne, Victoria, Australia), Nicholas Blackburn (Menzies Institute for Medical Research, University of Tasmania, Hobart, Tasmania, Australia), Bennet McComish (Menzies Institute for Medical Research, University of Tasmania, Hobart, Tasmania, Australia), Deborah Mason (New Zealand Brain Research Institute, Christchurch, New Zealand).

## DATA AVAILABILITY

ANZgene Consortium data is available under controlled access, and access requests can be made via the MS Australia secretariat using an official ANZgene Data Request form. GeneMSA data is available through dbGaP (accession number: phs000171.v1.p1). WTCCC2 genotyping data is available through the European Genome-phenome Archive (EGA; EGAS00000000101). Immune cell gene expression data is also available in the EGA under EGAS00001007254. Longitudinal MS cohort data is available under controlled access, and access requests with scientifically sound proposals can be made in writing to Associate Professor Vilija Jokubaitis or Professor Helmut Butzkueven.

