## Supplementary Figure S1-20 for "C4 genetic structural variations affect multiple sclerosis risk and progression"

### Figure S1. C4 diplotype associations with risk of MS onset and disability progression

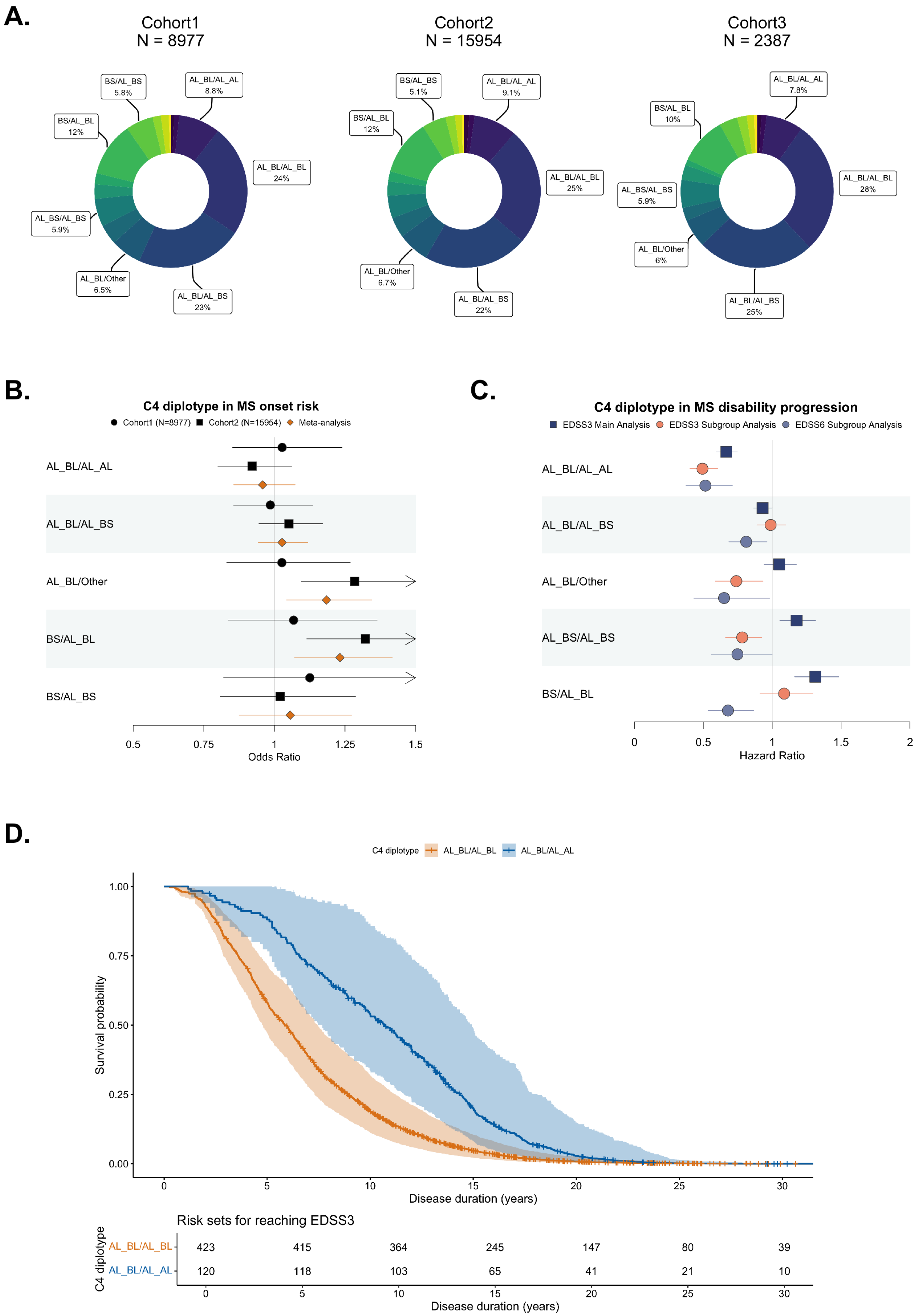

**Figure S2. Correlation of C4 copy numbers and C4 haplotypes with covariates included in multivariable models.**

A.

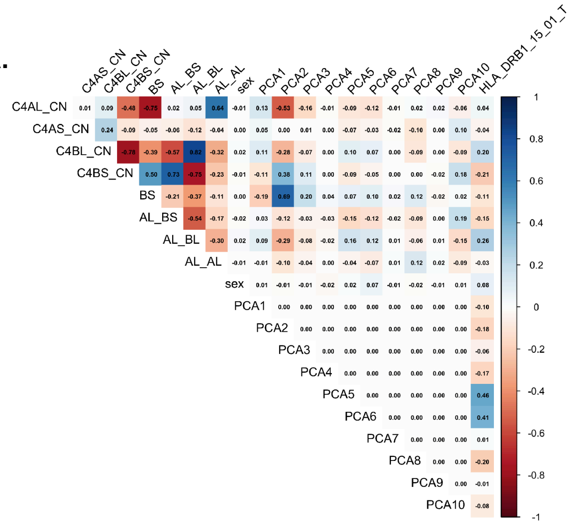


B.

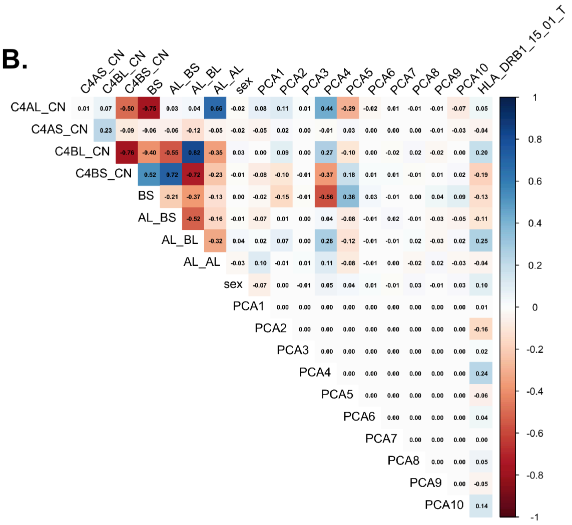


C.

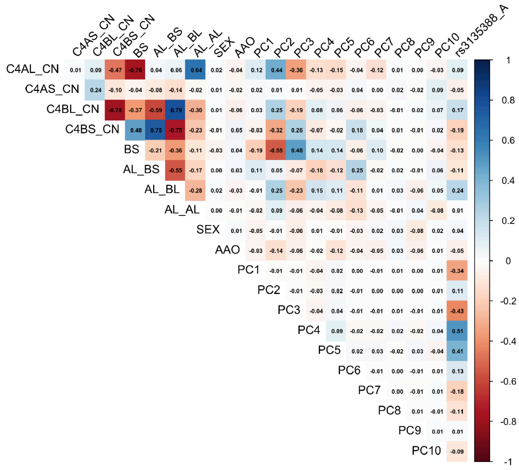

### Figure S3. Sex-stratified analysis of C4 associations with risk of MS onset and disability progression.

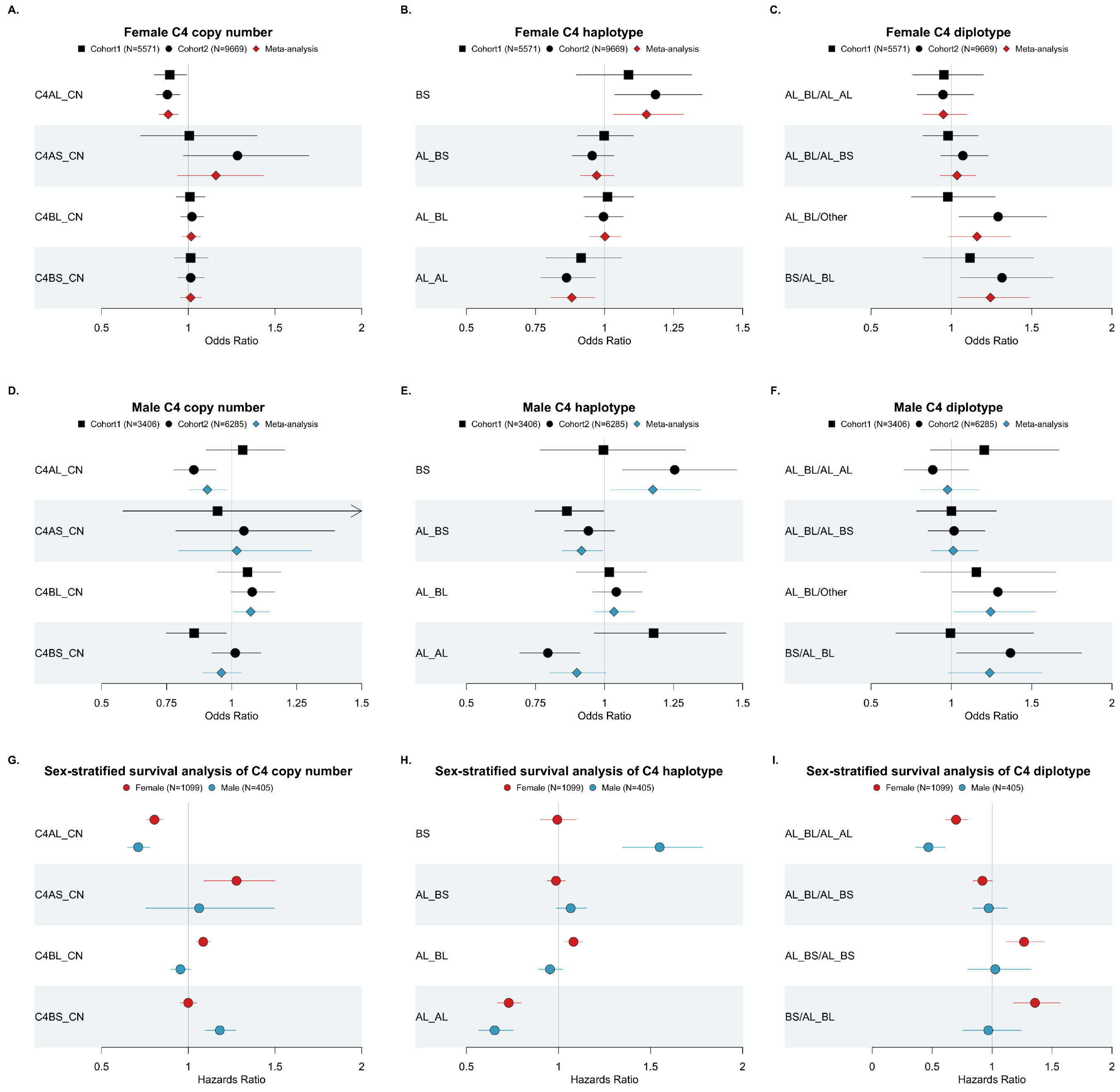

**Figure S4. Immune cell-specific *C4* expression profiles by C4BL copy numbers.**

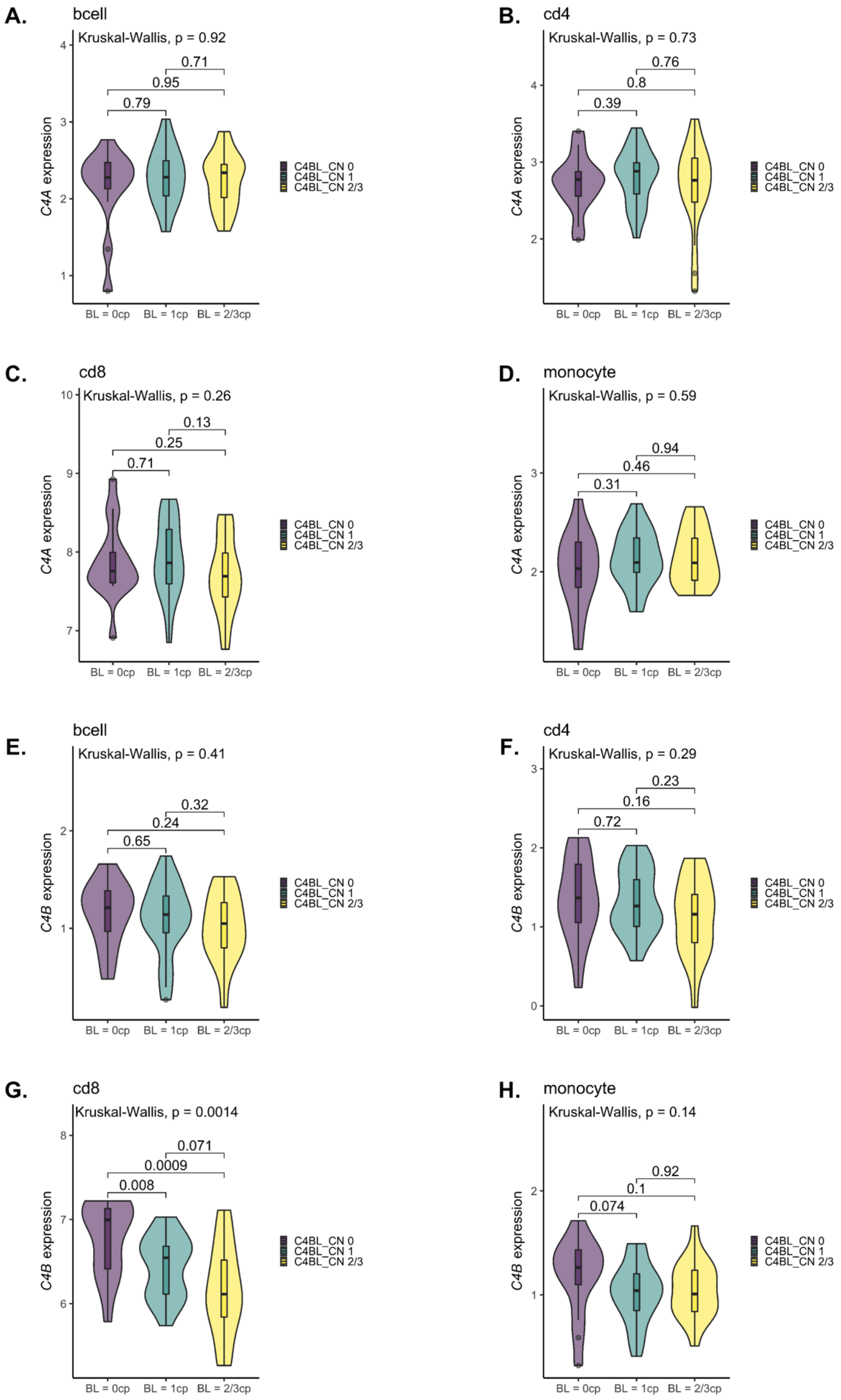

**Figure S5. Immune cell-specific *C4* expression profiles by C4BS copy numbers.**

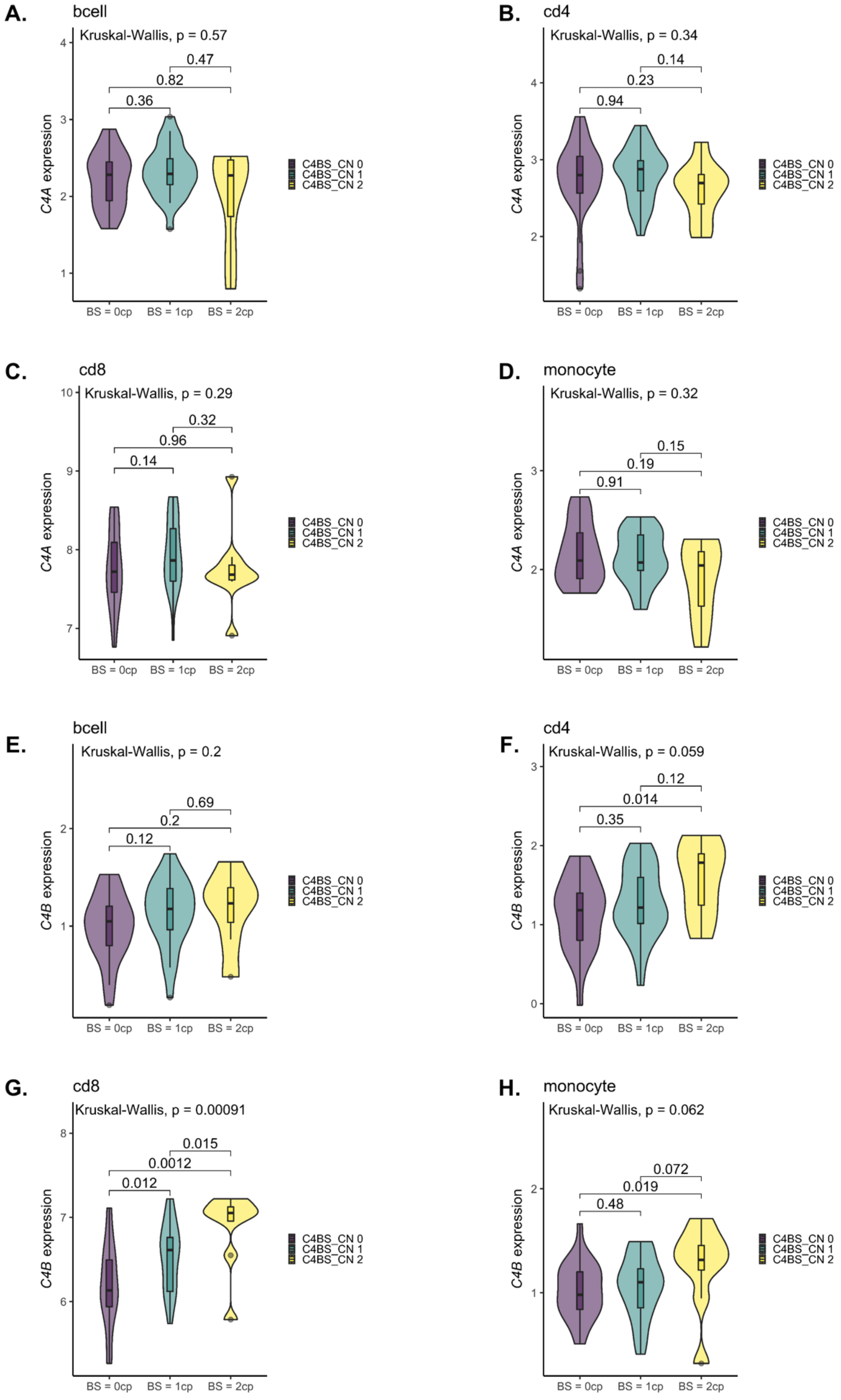

**Figure S6. Immune cell-specific *C4* expression profiles by C4AL copy numbers.**

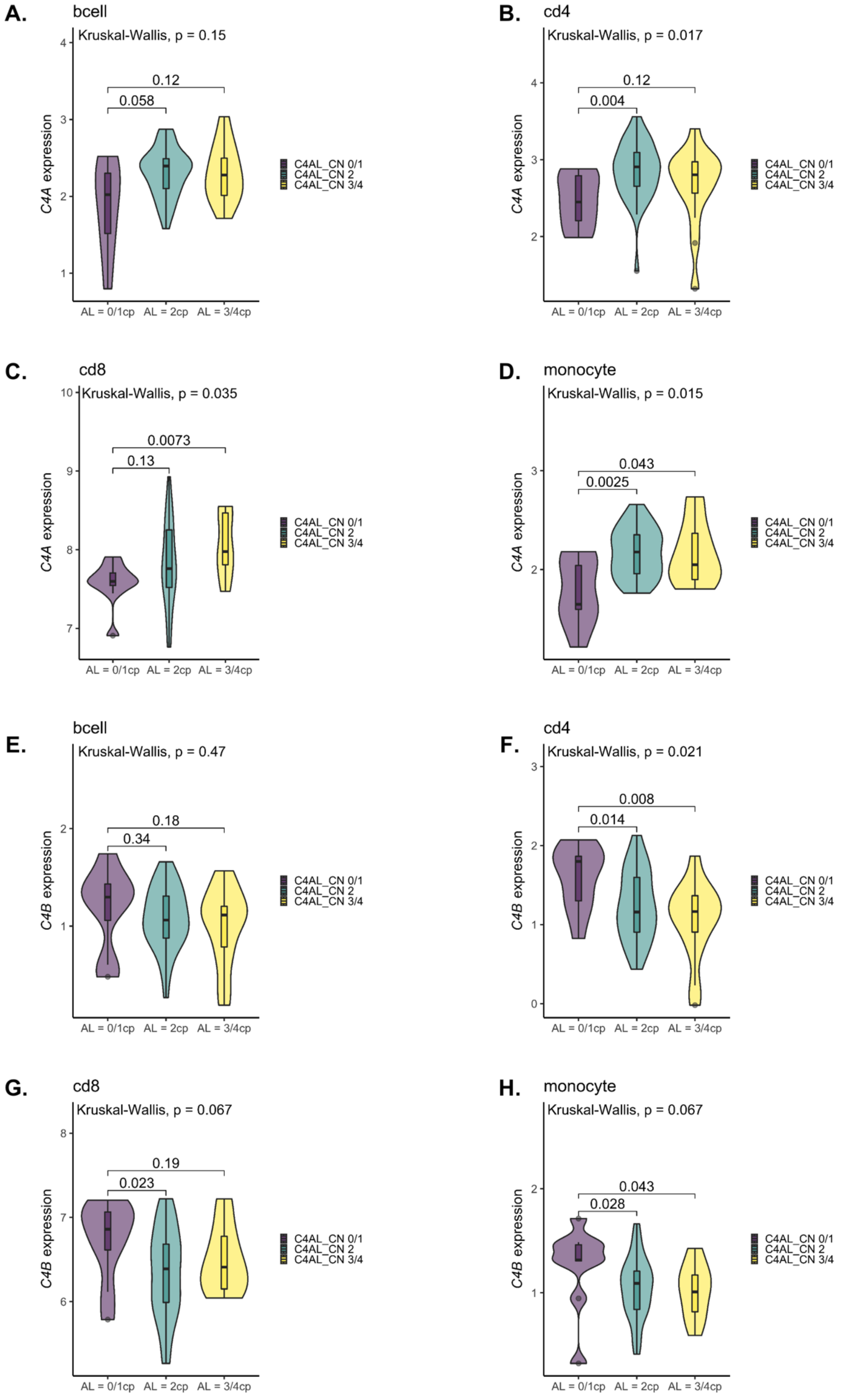

**Figure S7. Immune cell-specific *C4* expression profiles by AL_BL haplotype.**

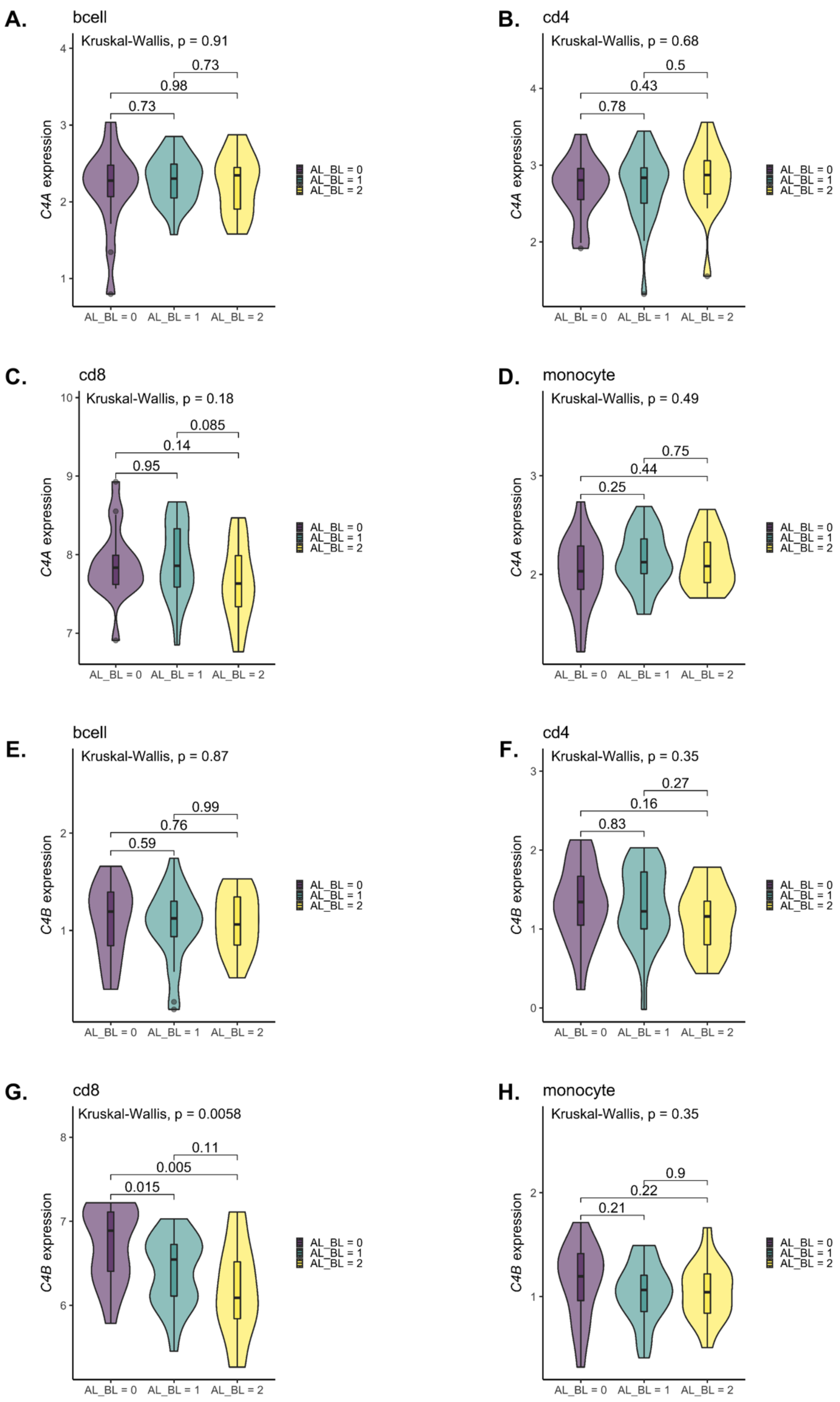

**Figure S8. Immune cell-specific *C4* expression profiles by AL_BS haplotype.**

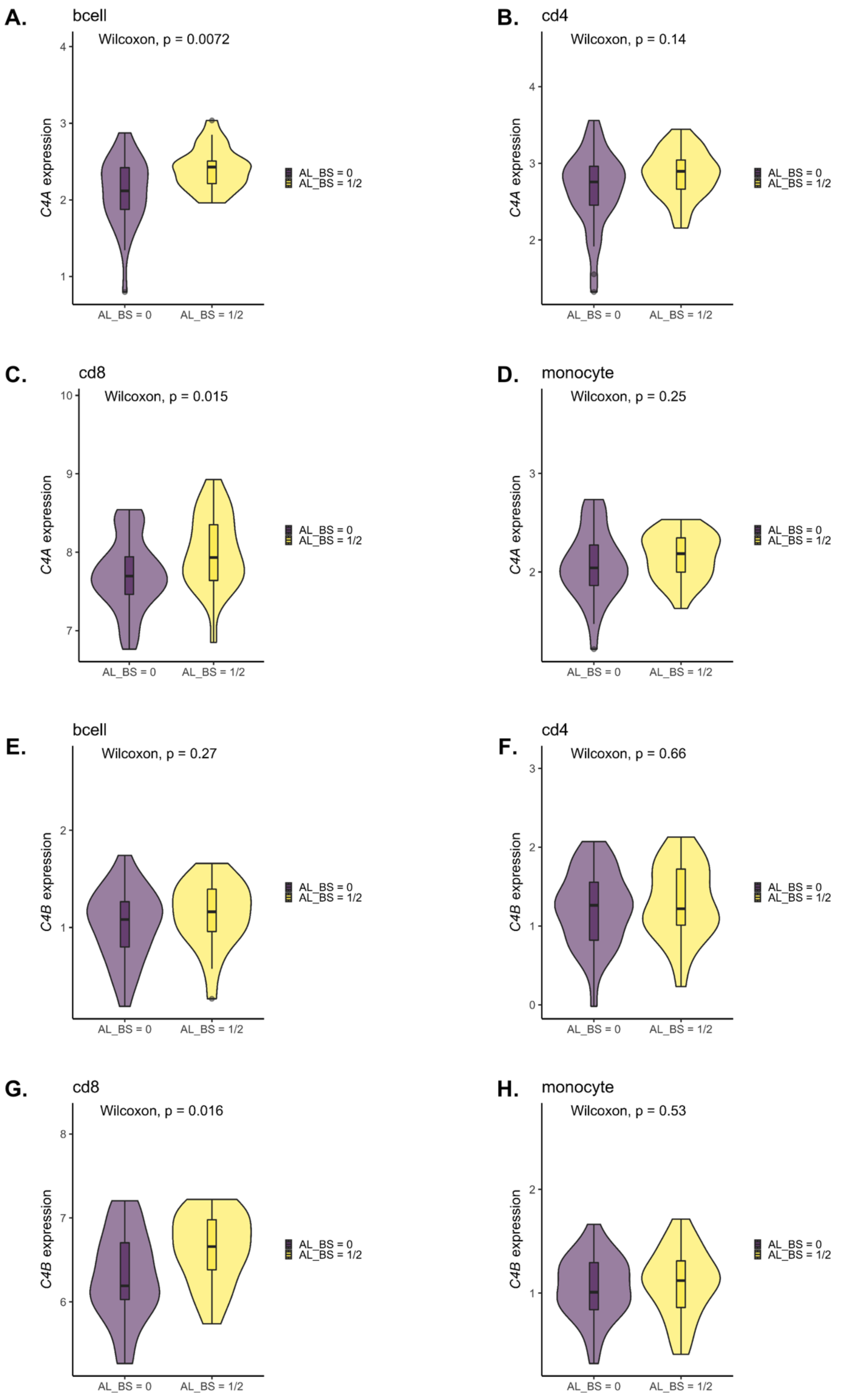

**Figure S9. Immune cell-specific *C4* expression profiles by AL_AL haplotype.**

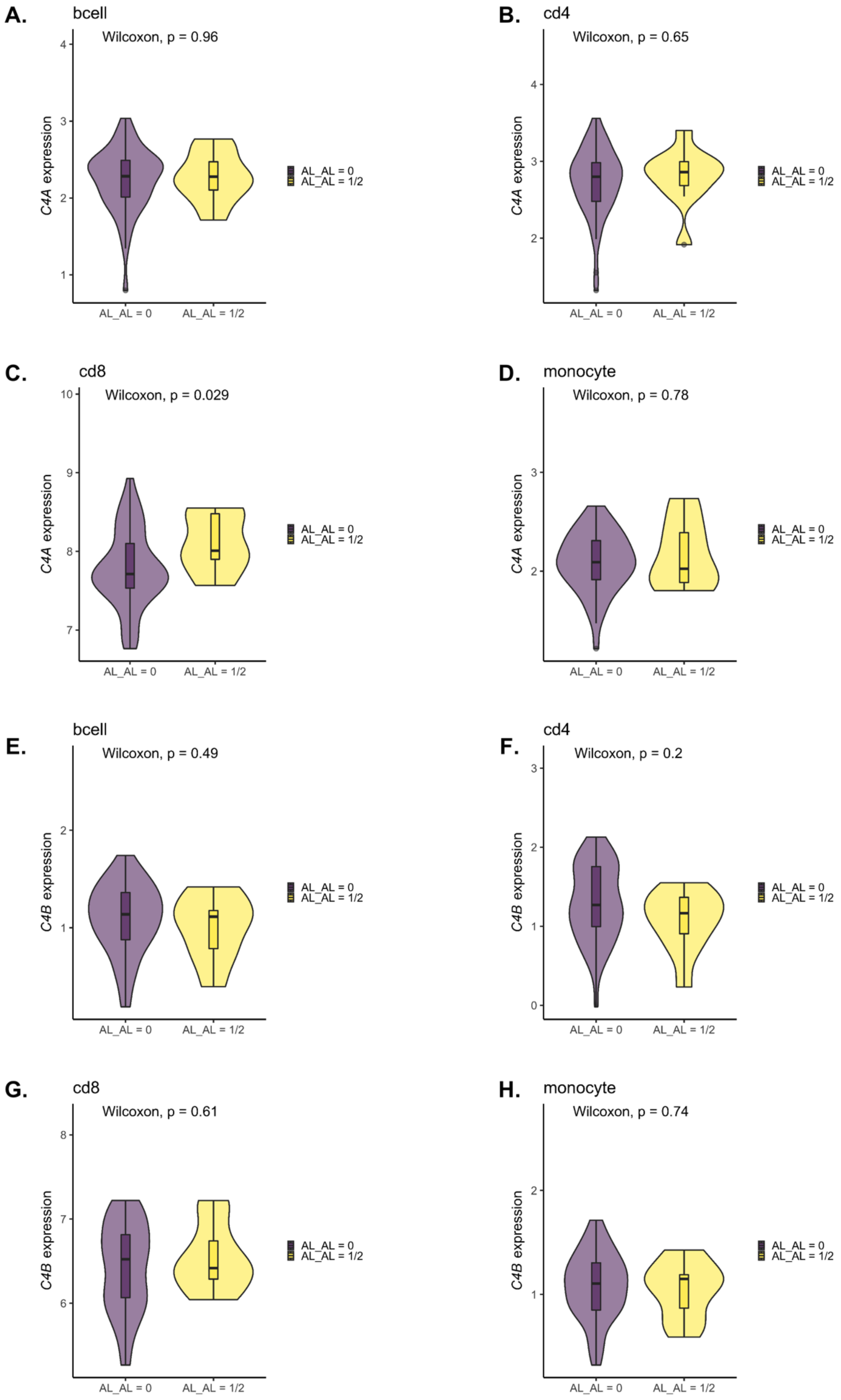

**Figure S10. Immune cell-specific *C4* expression profiles by BS haplotype.**

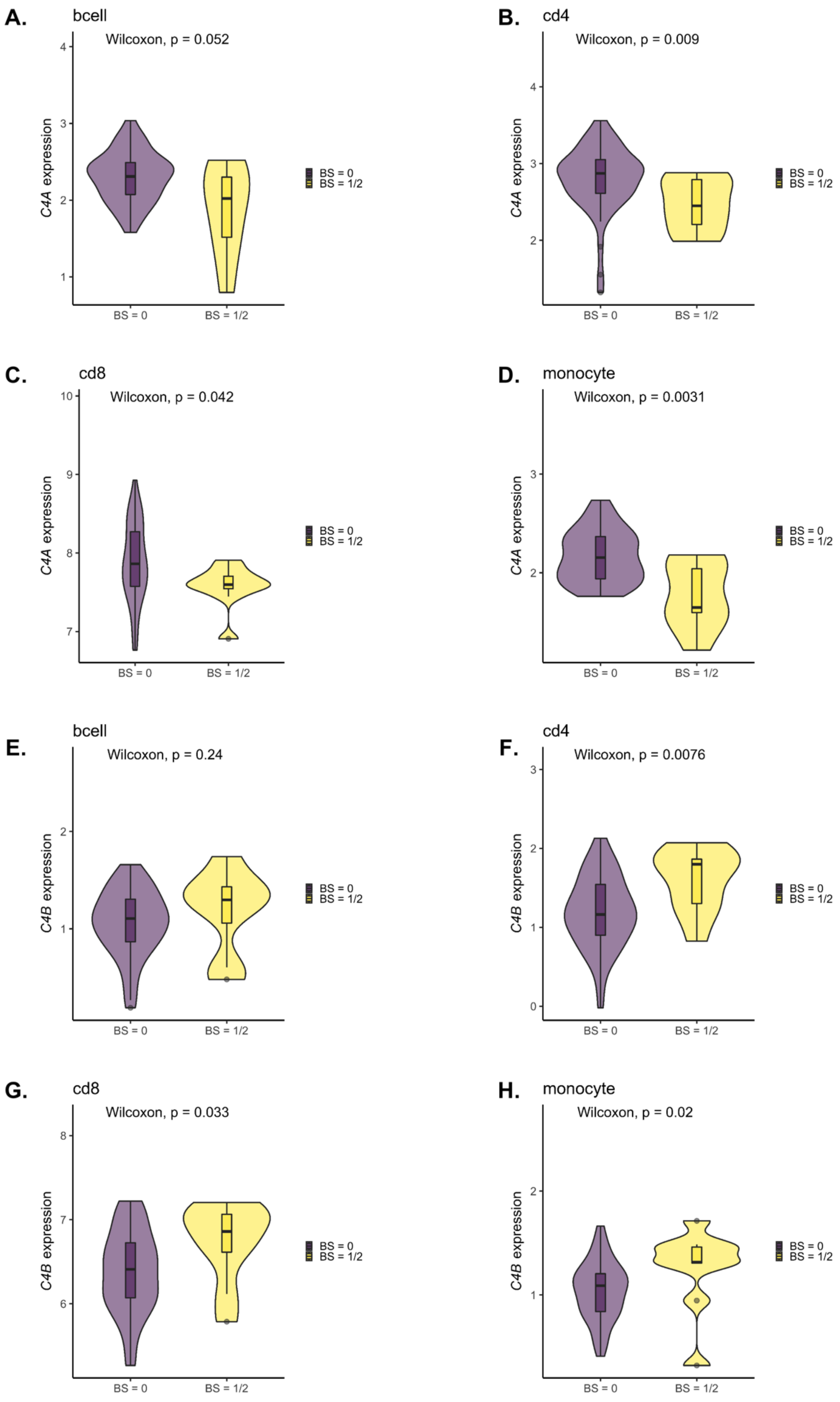

**Figure S11. Immune cell-specific *C4* expression profiles by C4AL copy numbers and case-control status.**

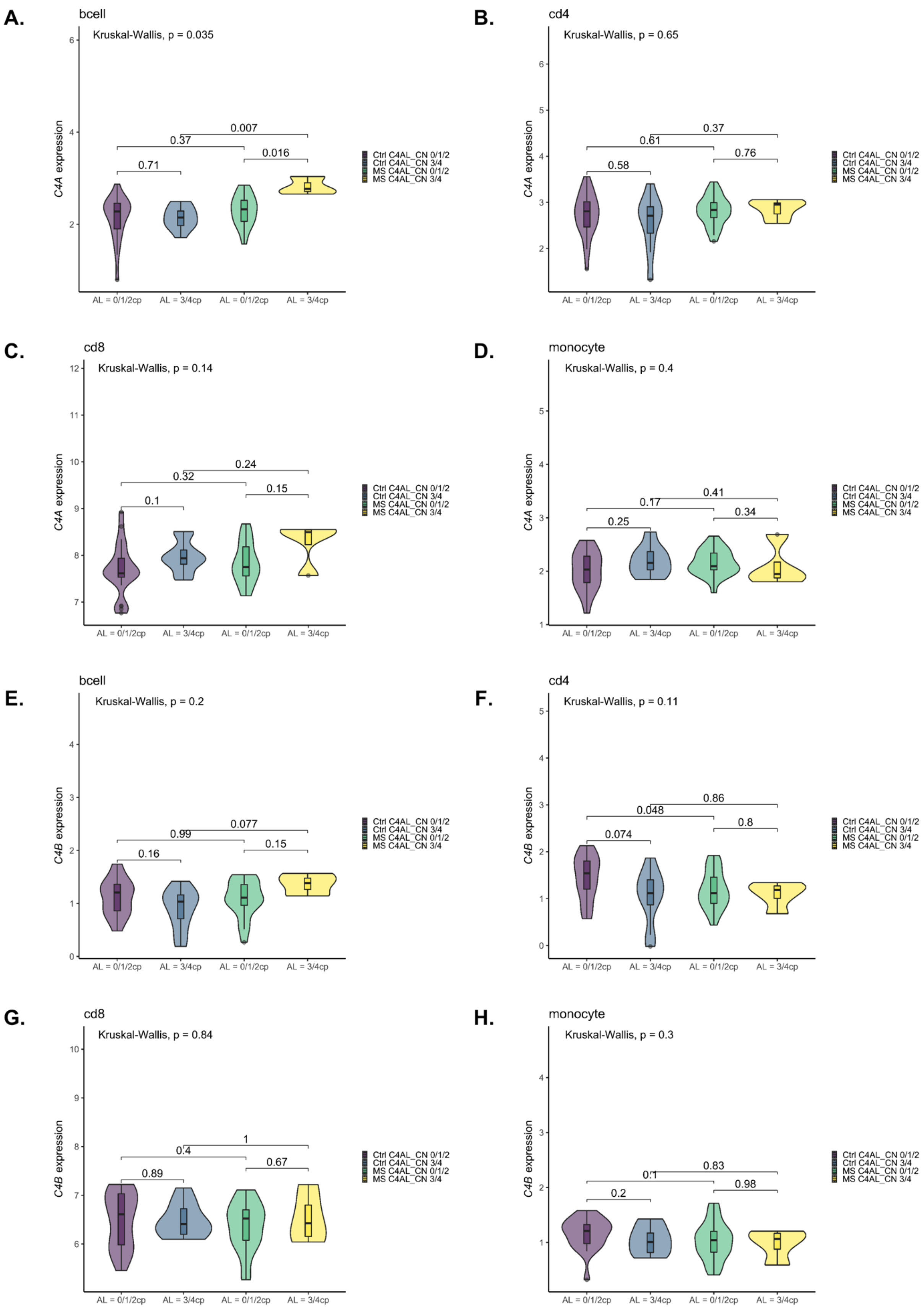

**Figure S12. Immune cell-specific *C4* expression profiles by C4BL copy numbers and case-control status.**

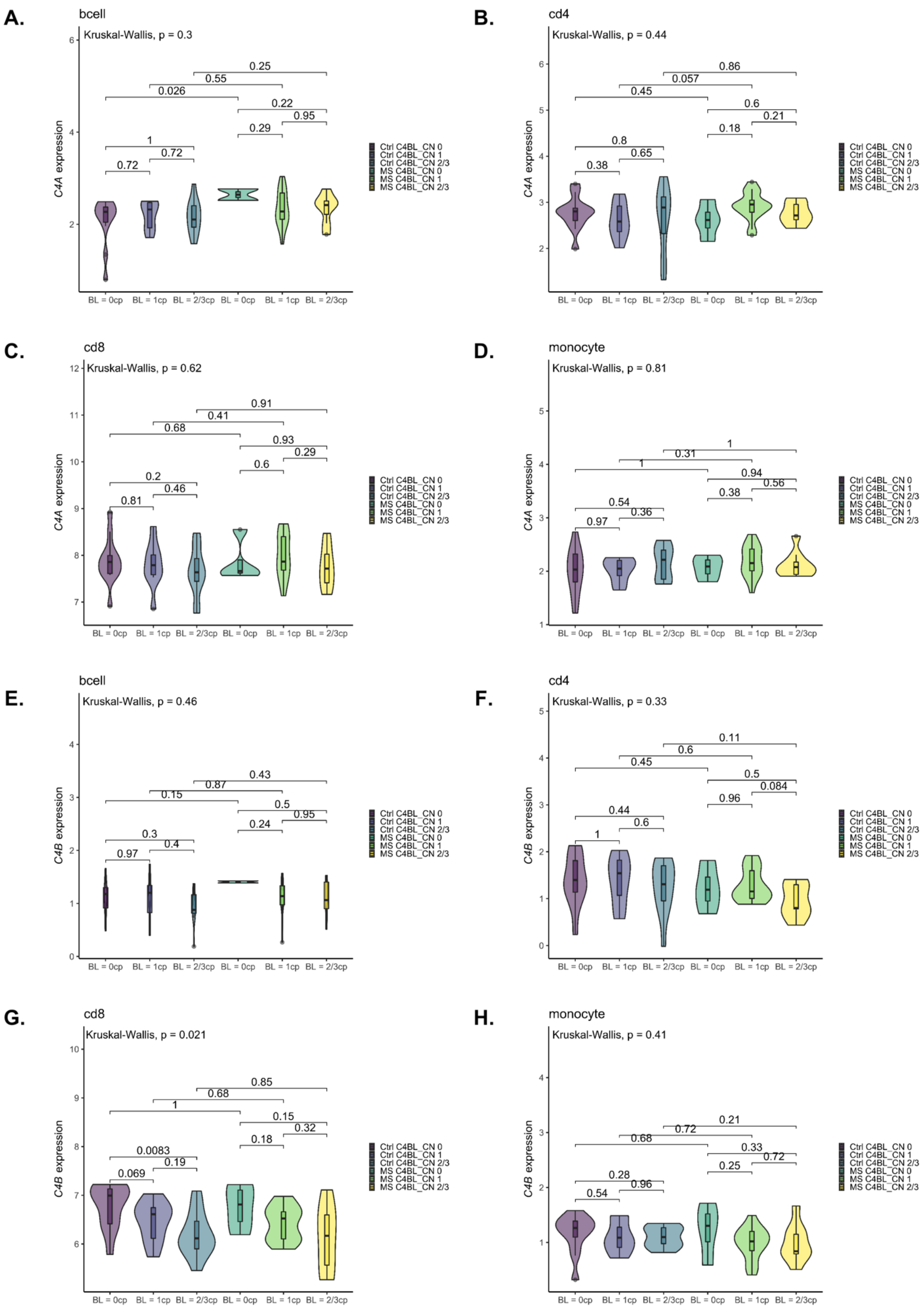

**Figure S13. Immune cell-specific *C4* expression profiles by C4BS copy numbers and case-control status.**

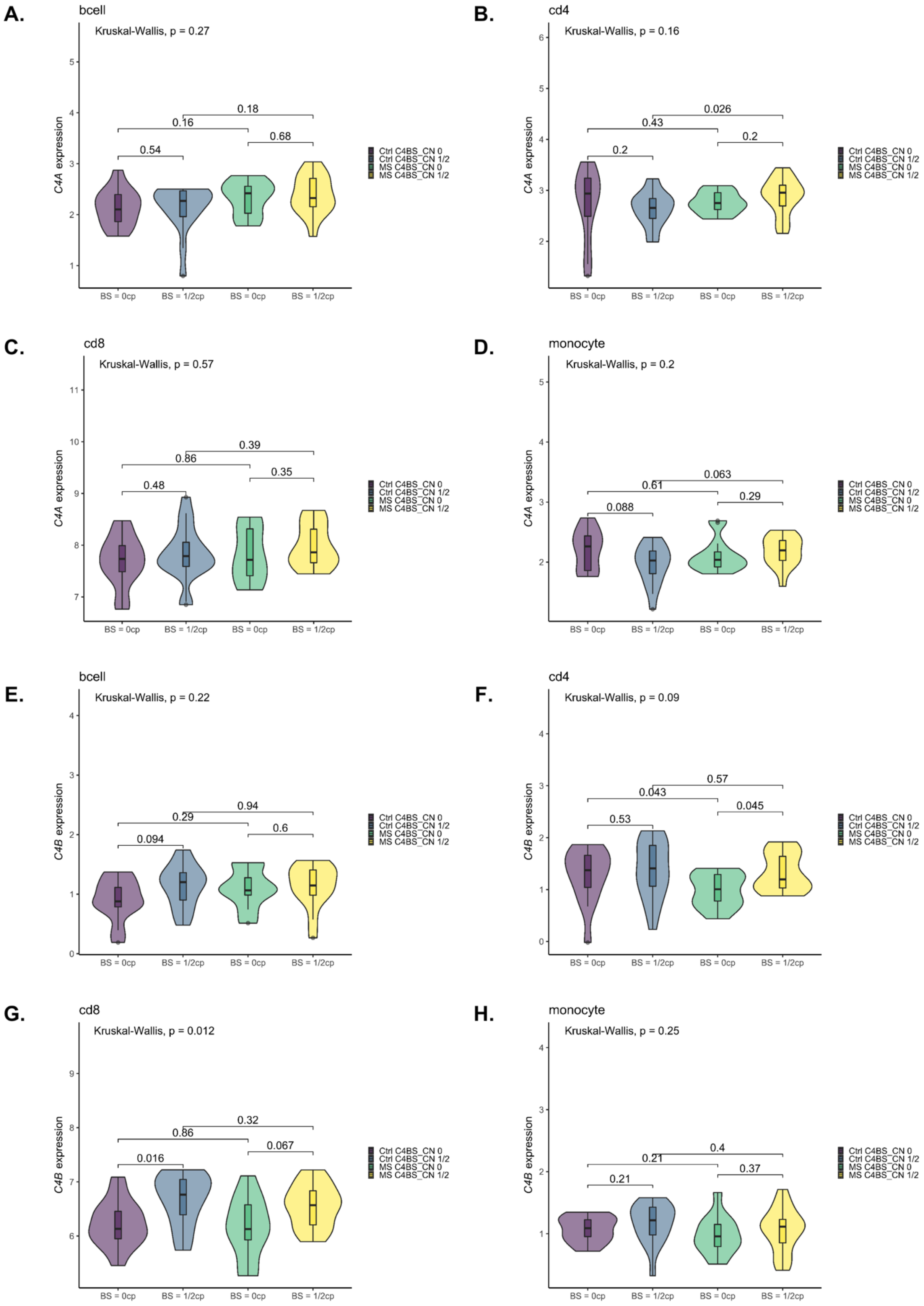

**Figure S14. Immune cell-specific *C4* expression profiles by AL_BL haplotype and case-control status.**

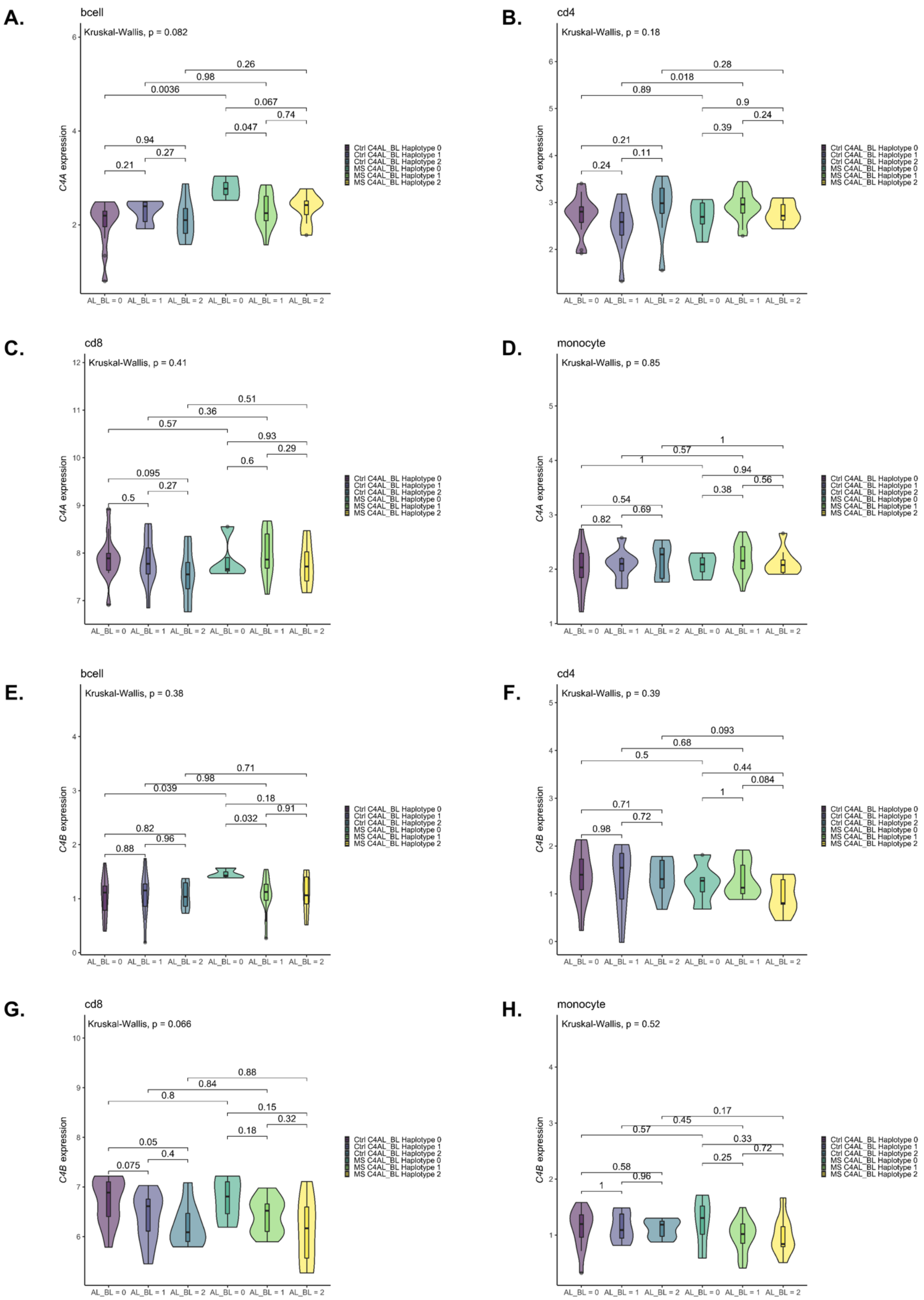

**Figure S15. Immune cell-specific *C4* expression profiles by AL_BS haplotype and case-control status.**

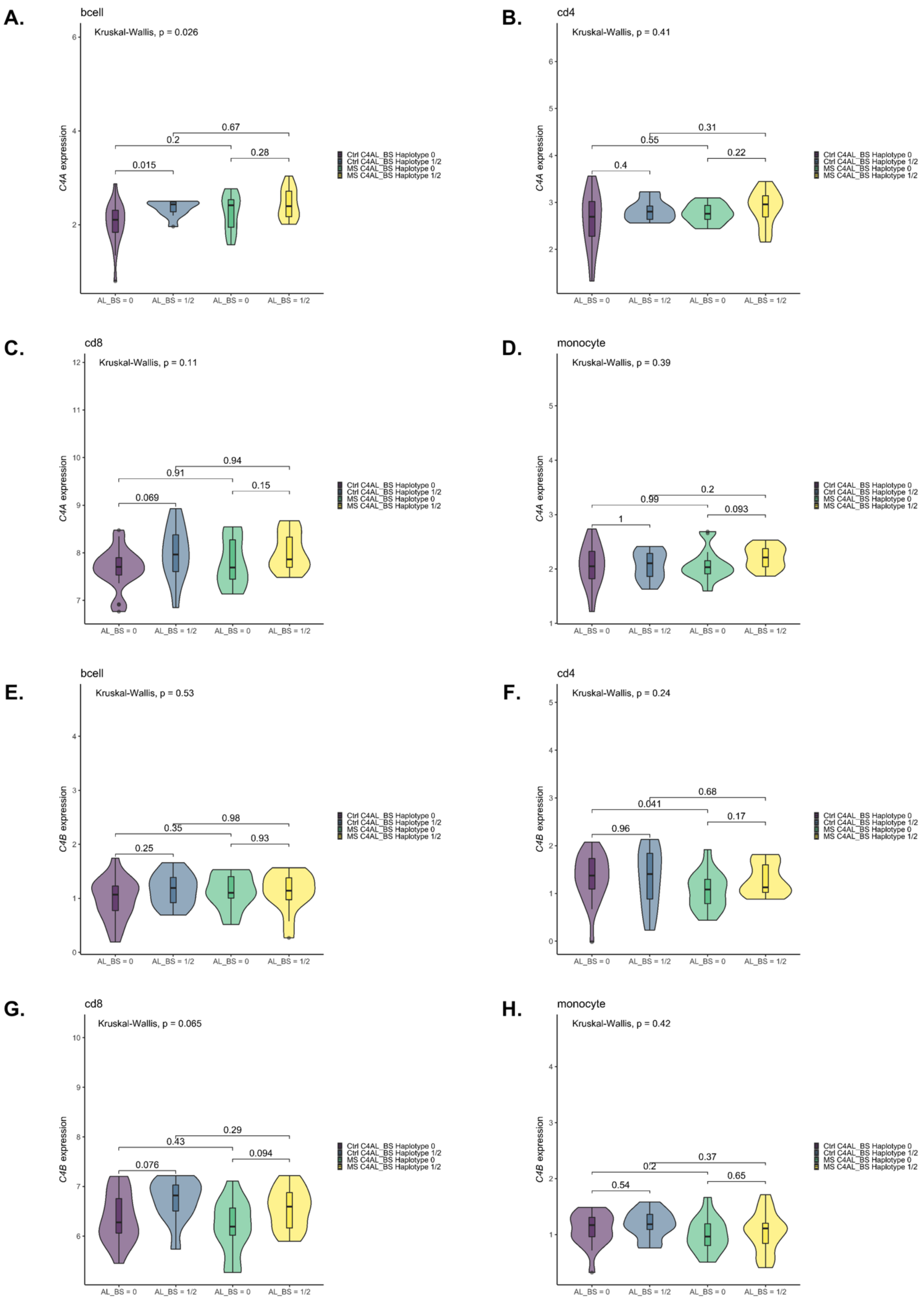

**Figure S16. Immune cell-specific *C4* expression profiles by AL_AL haplotype and case-control status.**

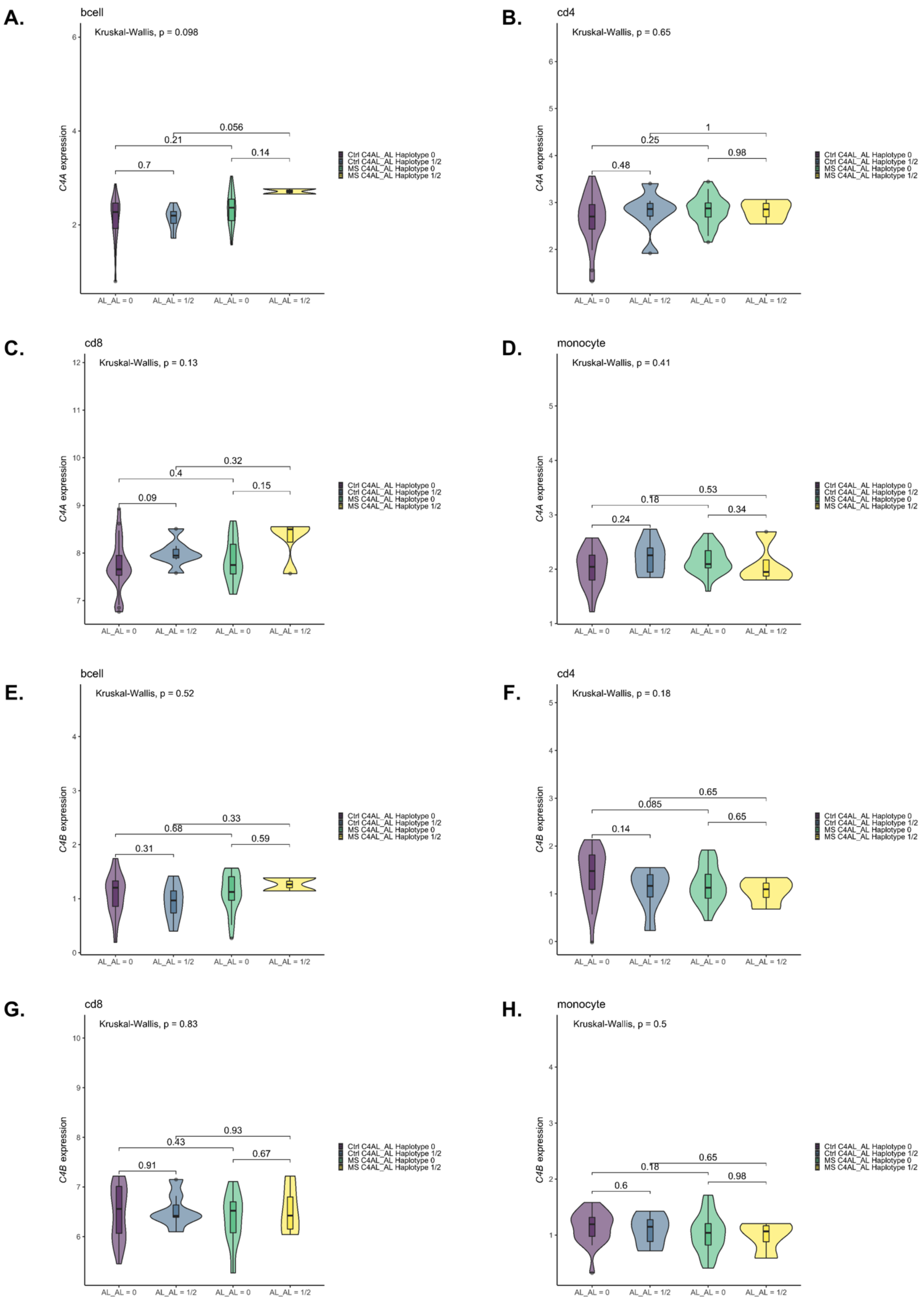

**Figure S17. Antigen processing and presentation pathway from KEGG pathway over-representation analysis.**

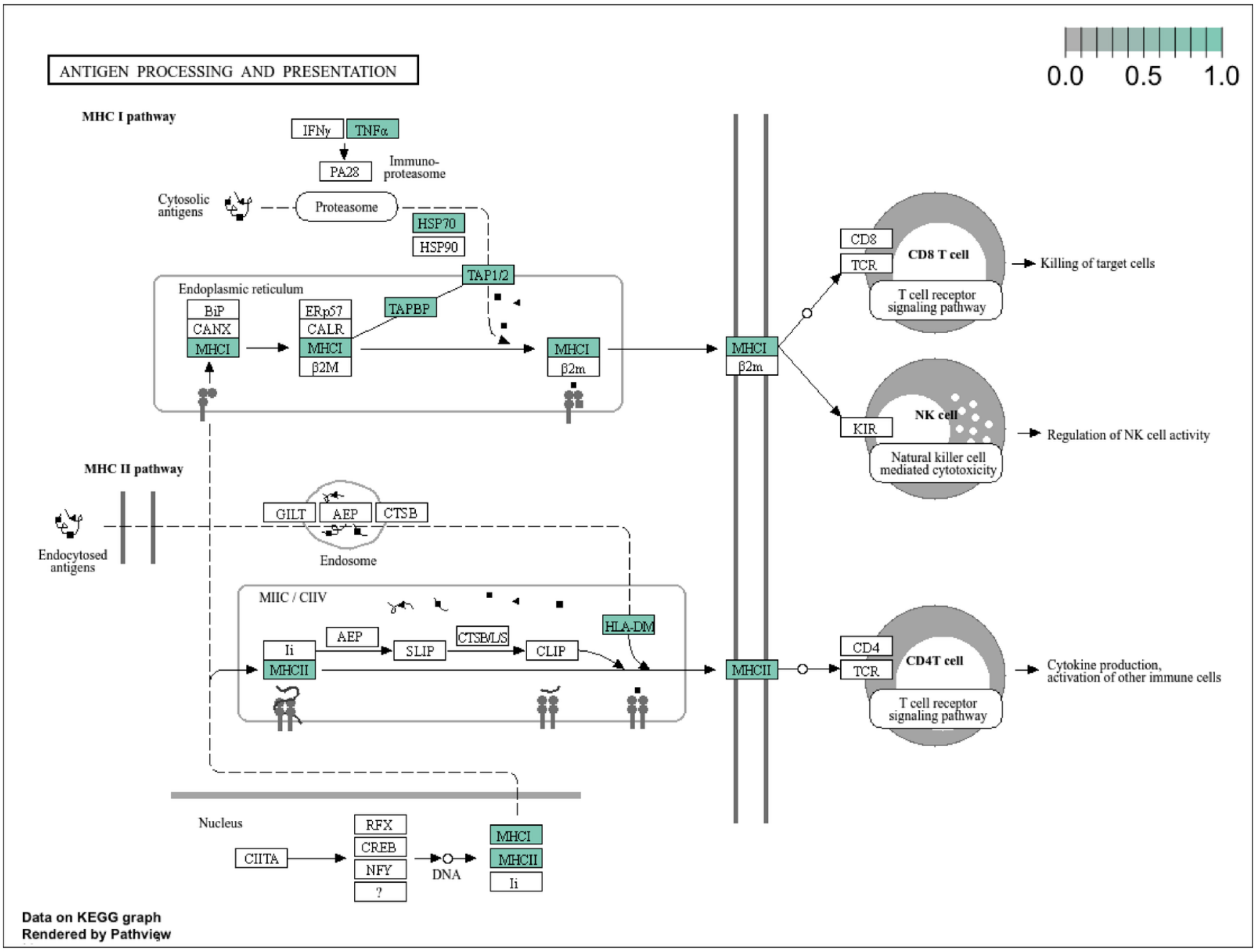

**Figure S18. Systemic lupus erythematosus pathway from KEGG pathway over-representation analysis.**

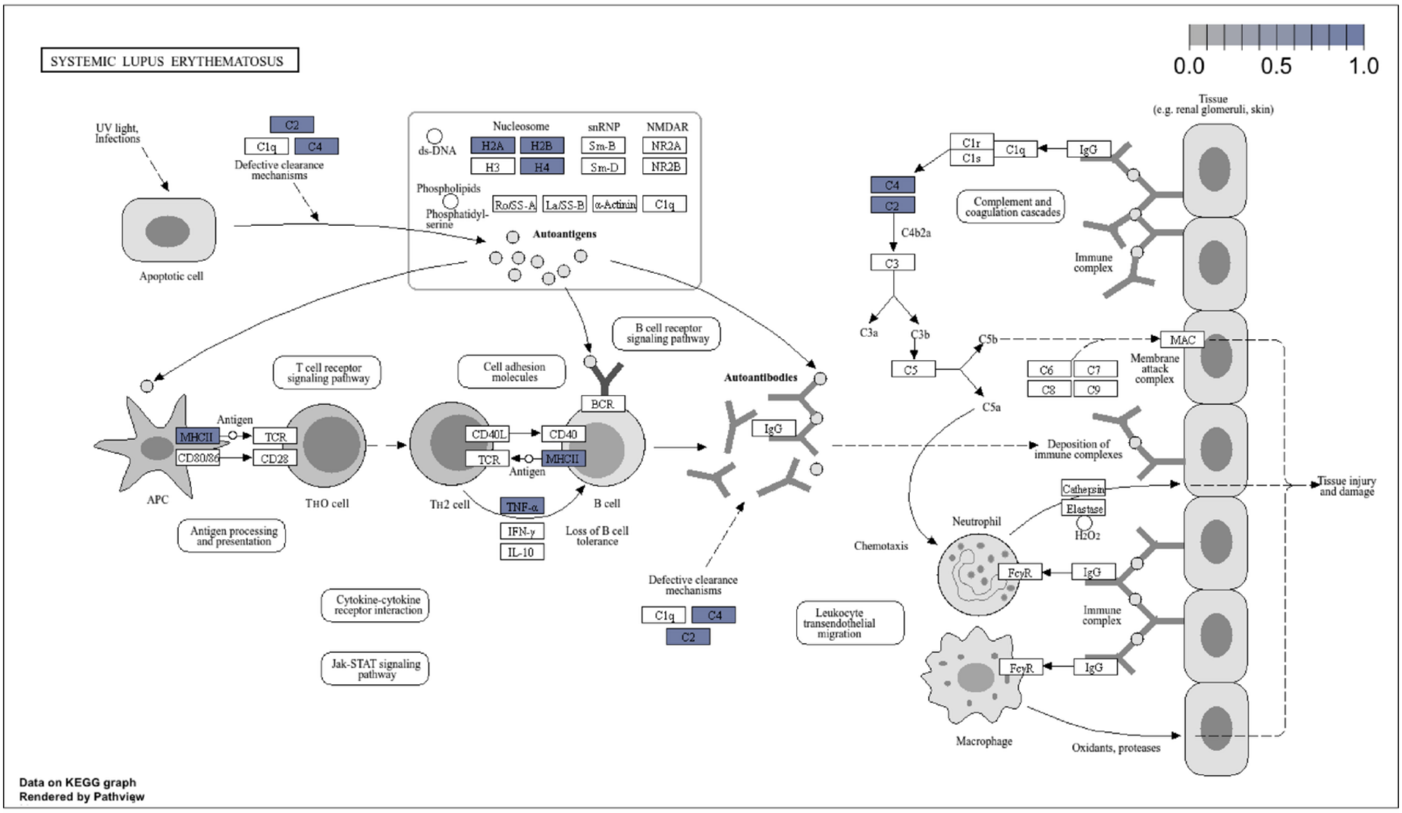

**Figure S19. Epstein-Barr virus infection pathway from KEGG pathway over-representation analysis.**

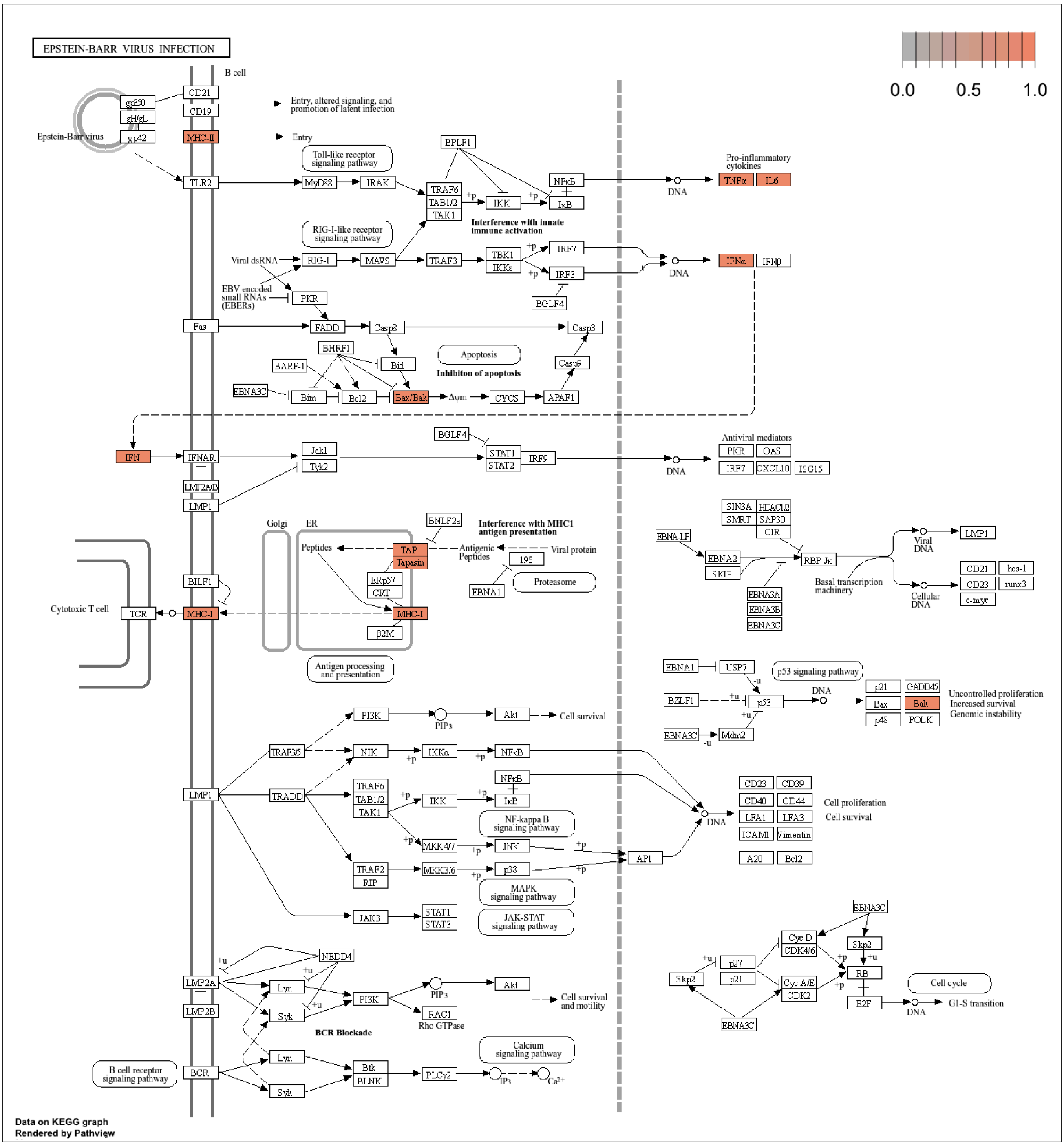

**Figure S20. Inflammatory bowel disease pathway from KEGG pathway over-representation analysis.**

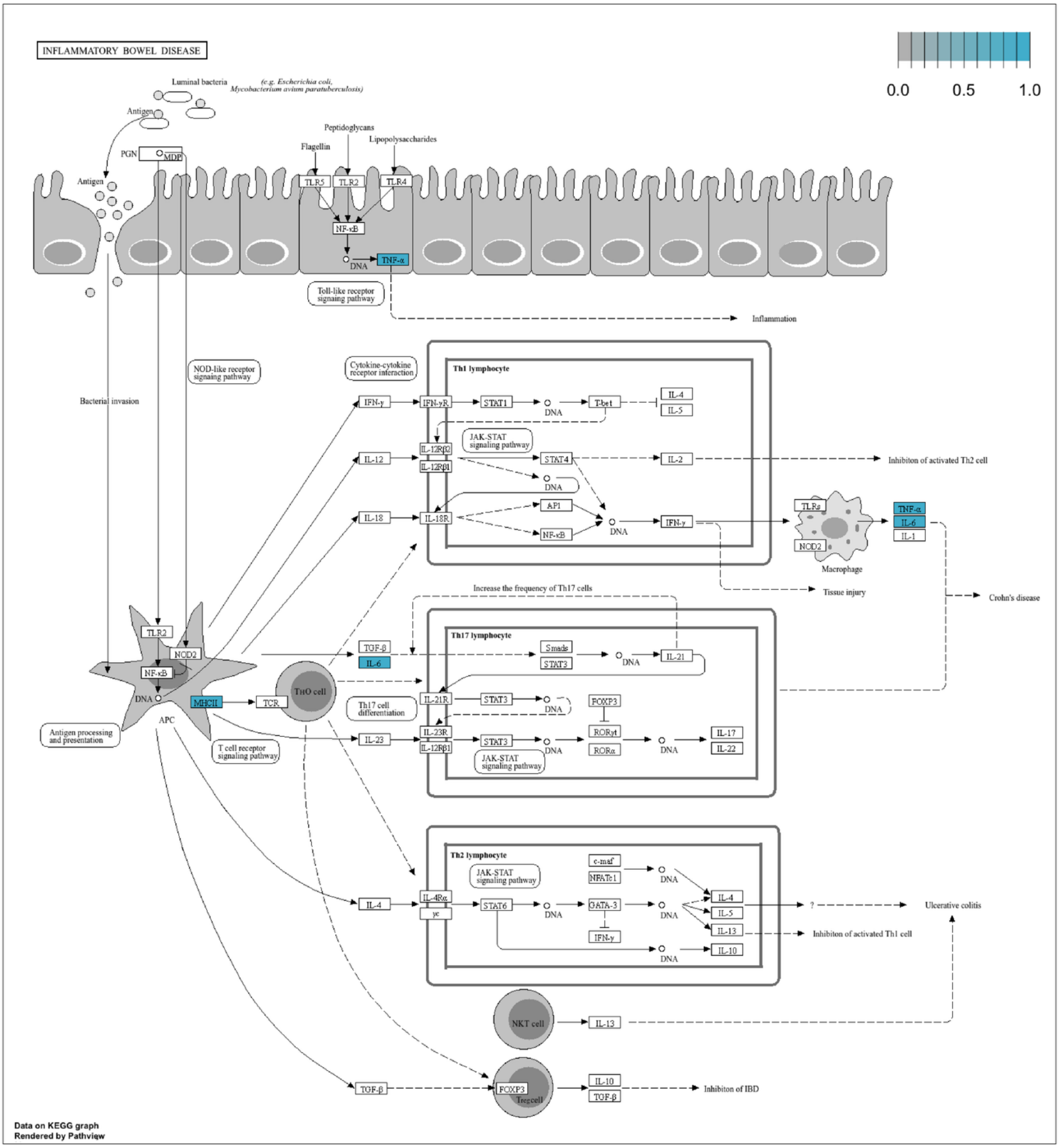
